# Secular trends of incidence and prevalence of parkinsonism and subtypes: A cohort study in the United Kingdom

**DOI:** 10.1101/2024.09.19.24313907

**Authors:** Xihang Chen, Nicola L. Barclay, Marta Pineda-Moncusí, Martí Català Sabaté, Laura Molina-Porcel, Wai Yi Man, Antonella Delmestri, Daniel Prieto-Alhambra, Annika M. Jödicke, Danielle Newby

## Abstract

**Background:** While summarized under the umbrella term “Parkinsonism”, several subtypes with different etiologies exist, including Parkinson’s Disease, Vascular Parkinsonism and Drug-induced Parkinsonism. However, evidence on their incidence and prevalence remains limited.

**Objectives:** To evaluate secular trends of incidence and prevalence of parkinsonism, Parkinson’s Disease, Vascular Parkinsonism, and Drug-induced Parkinsonism from 2007 to 2021 in the United Kingdom.

**Methods:** We used primary care data, Clinical Practice Research Datalink GOLD, from the United Kingdom. Individuals were included if they were registered from January 2007 to December 2021 with at least one year of prior observation. Incidence and prevalence were calculated on a yearly basis with 95% confidence intervals and then stratified by age and sex.

**Results:** From 2007 to 2019, the incidence of parkinsonism and Parkinson’s Disease decreased, with Parkinson’s Disease incidence dropping from 35.86 (95% confidence interval: 34.22 – 37.56) to 31.40 (29.40 – 33.50) per 100,000 person-years. The prevalence of parkinsonism and Parkinson’s Disease increased from 0.22% (0.22% - 0.23%) and 0.21% (0.21% - 0.22%) in 2007 to peak in 2016 with 0.25% (0.25% - 0.26%) and 0.23% (0.23% - 0.24%) respectively. The number of Vascular Parkinsonism diagnoses have increased from 2010, whereas incidence and prevalence of Drug-induced Parkinsonism remained stable. Incidence and prevalence increased with age and were generally higher in males, except for Drug-induced Parkinsonism, which was slightly higher in females.

**Conclusions:** Given its association with aging, parkinsonism and these subtypes continue to present an increasing challenge to our aging society.

## Introduction

Parkinsonism is an umbrella term used to describe a syndrome characterized by symptoms including muscle rigidity, tremors and slowness of movement [1]. These symptoms are commonly associated with Parkinson’s disease (PD), which is caused by the progressive degeneration and loss of dopamine-producing neurons in the brain, particularly in the substantia nigra [2]. PD is the second most common age-related neurodegenerative disease after Alzheimer’s Disease [2]. It not only poses a major burden on affected patients but is also becoming an increasingly significant financial burden on society [3].

Parkinsonism can also be caused by other conditions and factors, with other subtypes including Vascular Parkinsonism (VP) and Drug-induced Parkinsonism (DIP) as well as other progressive brain conditions [1]. VP is caused by ischemic cerebrovascular diseases which affect the brain areas involved in movement control, such as the basal ganglia, rather than a gradual loss of nerve cells observed in PD [4]. On the other hand, DIP is caused by medications that block dopamine receptors in the brain [5,6]. Drugs known to induce DIP include certain antipsychotics, antidepressants, and antiepileptics; however, symptoms can be reversible after withdrawal of the causative drug [5,6].

Critically, there is a lack of up-to-date evidence on the secular trends of the incidence and prevalence of parkinsonism and these subtypes, particularly in different sex and age groups. Studies have been carried out to investigate the incidence and prevalence of PD, however they did not consider parkinsonism and other subtypes of parkinsonism and have shown different temporal trends around the globe [7–10]. For example, a study in Norway has shown stable incidence and increasing prevalence [8] while a study in South Korea has shown increasing incidence and increasing prevalence [9]. Moreover, although other studies considered parkinsonism and subtypes, they were conducted regionally which may affect its generalizability [11,12]. Furthermore, although these two studies stratified results by sex and age groups, they did not consider temporal trends within each group.

In the light of this, a comprehensive assessment of disease burden over time is vital to better inform decisions regarding screening and prevention. Therefore, the aim of this study is to examine the secular trends of incidence and prevalence of parkinsonism, PD, VP and DIP from 2007 to 2021 in the UK for the whole population and stratified by age and sex.

## Methods

### Study design, setting, and data sources

This was a descriptive, population-based cohort study using routinely collected primary care data from the United Kingdom (UK). The database for this study was the Clinical Practice Research Datalink (CPRD) GOLD database. CPRD GOLD is an established database documenting the pseudo-anonymized patient-level data contributed by General Practitioners (GPs) across the UK, covering approximately 7% of the UK population [13]. CPRD GOLD was mapped to OMOP (Observational Medical Outcomes Partnership) CDM (Common Data Model) [14] to standardize analytics. The use of CPRD GOLD for this study was approved through CPRD’s Research Data Governance (RDG) Process (Protocol number: 22_002351).

### Study participants and time at risk

People were eligible for this study if they were aged above 18 with at least one year of prior observation registered in CPRD GOLD from 1st Jan 2007 to 31st Dec 2021. This formed the background population and was used to calculate time at risk in person-years. Individuals with a diagnosis of parkinsonism were included as the study population. They could also contribute to one or more subtype cohorts (PD, VP and/or DIP) when stratifying the analysis by these subtypes. However, not every individual with a diagnosis of parkinsonism needed to be part of the PD, VP, or DIP cohorts, as other subtypes, such as toxin-induced parkinsonism, were included to form the definition of parkinsonism.

Individuals were followed up to whichever came first: outcome of interest, practice stopped contributing to the database, patient left the practice, date of death, or the 31st of December 2021 (the end of the study period). For those diagnosed with either parkinsonism, PD or VP, each individual only contributed person-years once whereas for DIP, an individual could contribute person-years multiple times after one year washout from each DIP diagnosis.

### Outcome definitions

There were four study outcomes of interest: parkinsonism and three subtypes - PD, VP and DIP. Systematized Nomenclature of Medicine Clinical Terms (SNOMED CT) diagnostic codes were used for this study to identify diagnoses of parkinsonism, PD, VID and DIP using the R package CodelistGenerator [15]. The resulting computable phenotypes were then refined with the aid of R package CohortDiagnostics [16]. This package was used to identify additional codes of interest and to remove those highlighted as irrelevant based on feedback from clinicians with primary care and neurology expertise through an iterative process during the initial stages of analyses. The code lists used to define all the outcomes for this study can be found in Supplementary Table S1.

### Statistical methods

#### Baseline Characteristics

Baseline characteristics were summarized for patients with a diagnosis of parkinsonism, PD, VP, and DIP, respectively, using the R package PatientProfiles [17]. Median and IQR (interquartile range) were used for continuous variables whereas counts and percentages were used for categorical variables. The baseline characteristics included age, sex, conditions any time prior and medications within one year prior to the first onset of each study outcome.

#### Annual Crude Incidence Rates

For each year of the study period, a crude incidence rate was calculated for each outcome. For each incidence calculation, an individual can contribute at most once. Moreover, if an outcome of interest took place prior to 1^st^ Jan 2007 then this individual can no longer contribute time at risk. The incidence was calculated as a fraction between the number of incident cases and time at risk in person-years contributed by the background population that year. The rates calculated were then expressed as per 100,000 person-years with 95% confidence intervals (CIs).

#### Annual Crude Period Prevalence

Annual period prevalence was calculated for each outcome and expressed as percentages. The denominator was the number of people in the background population in the year of interest whereas the numerator was the number of cases of each outcome that year with 95% CIs calculated.

#### Incidence and Prevalence Stratification

All results were stratified by sex and by age. The age bands for the stratification were in 10 years except the first (18-30) and the last age band (80+). To protect patient privacy, we do not report results with fewer than five cases.

#### Annual Incidence and Prevalence Age Standardization

Age-standardized incidence rates and prevalence overall and by sex were calculated using 2013 European Standardized Population (ESP2013) [18]. ESP2013 serves as a standard age distribution to account for the change in age structure throughout the study period, and thus offers an age-standardized perspective on secular trends. The age groups in ESP2013 were merged and scaled so that the age groups were in ten-years except the first one which was 18-29.

All incidence and prevalence analyses were performed using R (version 4.2.3) and the R package IncidencePrevalence [19]. The analytical code for this study can be accessed via https://github.com/oxford-pharmacoepi/ParkinsonsIncidencePrevalence.

## Results

### Baseline Characteristics

Out of a total of 17,054,819 people available in CPRD GOLD from 1st Jan 2007 to 31st Dec 2021, 9,604,592 were eligible for this study. The attrition tables for each analysis can be found in Supplementary Table S2. There was a total of 21,680, 20,006, 1126 and 809 patients with a diagnosis of Parkinsonism, PD, VP and DIP, respectively, across the study period (Table 1).

**Table 1:** Baseline characteristics of patients with a diagnosis of parkinsonism and the subtypes PD, VP, and DIP between 2007 and 2021.

| Characteristic | Parkinsonism | PD | VP | DIP |
| --- | --- | --- | --- | --- |
| **Number of patients** | 21680 | 20006 | 1129 | 809 |
| **Sex: Female, n (%)** | 8516 (39.3) | 7793 (39) | 394 (39) | 450 (55.6) |
| **Age: Median (IQR)** | 76 (68-81) | 75 (68-81) | 80 (75-84) | 71 (63-80) |
| **Age Bands: n (%)** |  |  |  |  |
| 18 to 30 | 21 (0.1) | 12 (0.1) | 0 (0) | 9 (1.1) |
| 31 to 40 | 69 (0.3) | 56 (0.3) | 0 (0) | 9 (1.1) |
| 41 to 50 | 391 (1.8) | 356 (1.8) | <5 | 33 (4.1) |
| 51 to 60 | 1581 (7.3) | 1473 (7.4) | 15 (1.3) | 102 (12.6) |
| 61 to 70 | 4694 (21.7) | 4412 (22.1) | 107 (9.5) | 225 (27.8) |
| 71 to 80 | 8685 (40.1) | 8083 (40.4) | 478 (42.3) | 245 (30.3) |
| 81 to 90 | 5707 (26.3) | 5146 (25.7) | 477 (42.2) | 170 (21) |
| 91 to 100 | 530 (2.4) | 467 (2.3) | 50 (4.4) | 15 (1.9) |
| >100 | <5 | <5 | 0 (0) | <5 |
| **General Conditions (any time prior): n (%)** |  |  |  |  |
| Pneumonia | 631 (2.9) | 558 (2.8) | 62 (5.5) | 42 (5.2) |
| Gastroesophageal reflux disease | 729 (3.4) | 666 (3.3) | 58 (5.1) | 25 (3.1) |
| Chronic kidney disease | 3921 (18.1) | 3507 (17.5) | 329 (29.1) | 181 (22.4) |
| Stroke | 1044 (4.8) | 841 (4.2) | 189 (16.7) | 59 (7.3) |
| Anxiety | 3741 (17.3) | 3392 (17) | 194 (17.2) | 237 (29.3) |
| Dementia | 1817 (8.4) | 1582 (7.9) | 188 (16.7) | 91 (11.2) |
| Cardiovascular disease* | 7753 (35.8) | 6911 (34.5) | 724 (64.1) | 274 (33.9) |
| Hyperlipidemia | 2372 (10.9) | 2187 (10.9) | 137 (12.1) | 99 (12.2) |
| Diabetes | 2631 (12.1) | 2346 (11.7) | 226 (20) | 130 (16.1) |
| Myocardial infarction | 783 (3.6) | 683 (3.4) | 87 (7.7) | 24 (3) |
| Venous thromboembolism | 1070 (4.9) | 972 (4.9) | 75 (6.6) | 43 (5.3) |
| Malignant neoplastic disease | 3035 (14) | 2754 (13.8) | 224 (19.8) | 100 (12.4) |
| Chronic obstructive pulmonary disease | 1130 (5.2) | 1018 (5.1) | 101 (8.9) | 47 (5.8) |
| Heart failure | 796 (3.7) | 706 (3.5) | 80 (7.1) | 24 (3) |
| Osteoporosis | 1367 (6.3) | 1218 (6.1) | 120 (10.6) | 46 (5.7) |
| Asthma | 1757 (8.1) | 1628 (8.1) | 96 (8.5) | 69 (8.5) |
| Depressive disorder | 3507 (16.2) | 3094 (15.5) | 228 (20.2) | 273 (33.7) |
| Hypertension | 6407 (29.6) | 5902 (29.5) | 434 (38.4) | 198 (24.5) |
| **Medications (one year prior): n (%)** |  |  |  |  |
| Psycholeptics | 5638 (26) | 4790 (23.9) | 288 (25.5) | 688 (85) |
| Diuretics | 6066 (28) | 5520 (27.6) | 436 (38.6) | 222 (27.4) |
| Opioids | 6598 (30.4) | 6049 (30.2) | 391 (34.6) | 274 (33.9) |
| Antibacterials systemic | 8958 (41.3) | 8085 (40.4) | 608 (53.9) | 415 (51.3) |
| Systemic corticosteroids | 1944 (9) | 1783 (8.9) | 117 (10.4) | 88 (10.9) |
| Beta blocking agents | 6038 (27.9) | 5560 (27.8) | 342 (30.3) | 200 (24.7) |
| Drugs for obstructive airway disease | 4393 (20.3) | 4049 (20.2) | 257 (22.8) | 167 (20.6) |
| Calcium channel blockers | 5096 (23.5) | 4685 (23.4) | 322 (28.5) | 169 (20.9) |
| Anti-inflammatory antirheumatic | 5010 (23.1) | 4682 (23.4) | 212 (18.8) | 191 (23.6) |
| Renin-angiotensin-system (RAS)-acting agents | 7544 (34.8) | 6918 (34.6) | 524 (46.4) | 221 (27.3) |
| Antiepileptics | 2482 (11.4) | 2103 (10.5) | 173 (15.3) | 271 (33.5) |
| Drugs for acid related disorder | 8817 (40.7) | 8006 (40) | 566 (50.1) | 387 (47.8) |
| Antidepressants | 7153 (33) | 6365 (31.8) | 446 (39.5) | 468 (57.8) |
| Antithrombotics | 3818 (17.6) | 3344 (16.7) | 440 (39) | 136 (16.8) |
| Drugs used in diabetes | 2539 (11.7) | 2262 (11.3) | 208 (18.4) | 125 (15.5) |
| Lipid modifying agents | 9750 (45) | 8810 (44) | 711 (63) | 390 (48.2) |

IQR: Interquartile range; PD: Parkinson's Disease; VP: Vascular Parkinsonism; DIP: Drug-induced Parkinsonism
\*Cardiovascular diseases included venous thrombosis, pulmonary embolism, peripheral vascular disease, ischemic heart disease, heart failure, heart disease, coronary arteriosclerosis, cerebrovascular disease, and atrial fibrillation

Table 1 shows that parkinsonism, PD and VP patients were more common in males (M: ∼60%) whereas for DIP it was more common in females (F: 55.6%). Those diagnosed with VP had the highest median age (80 years, IQR: 75-84) whereas the median age for DIP was the lowest (71 years, IQR: 63-80). This was also reflected in their respective age group composition; for example, the percentage of VP patients aged above 80 was 46.6% whereas the same percentage for each parkinsonism, PD, and DIP was no greater than 30%.

Patients diagnosed with VP were more likely to have chronic kidney disease (CKD), dementia, myocardial infarction, stroke, hypertension, and cardiovascular disease with more prescriptions for diuretics, antithrombotics, RAS-acting agents and lipid modifying agents within one year prior to the first onset compared to parkinsonism, PD and DIP. Similarly, patients diagnosed with DIP were more likely to be diagnosed with anxiety and depressive disorder, and to be prescribed with antidepressants, antiepileptics and psycholeptics compared to parkinsonism, PD and VP.

### Crude Incidence

The annual incidence of parkinsonism and PD decreased slightly from 2007 to 2019, with the incidence of PD being 35.86 per 100,000 person-years (pys) (95% CI: 34.22 – 37.56) and 31.40 per 100,000 pys (29.40 – 33.50) in 2007 and 2019 respectively (Figure 1). In 2020 and 2021, during the COVID-19 pandemic, this incidence dipped to 22.90 (21.13 – 24.76) and 28.72 (26.64 – 30.93). The incidence of VP increased from 0.17 (0.08 – 0.33) in 2009 to 2.03 in 2021 (1.51 – 2.68), peaking in 2019 with 3.18 per 100,000 pys (2.57 – 3.90). The incidence of DIP fluctuated between 0.9 to 1.6 per 100,000 pys across the whole study period.

**Figure 1:**
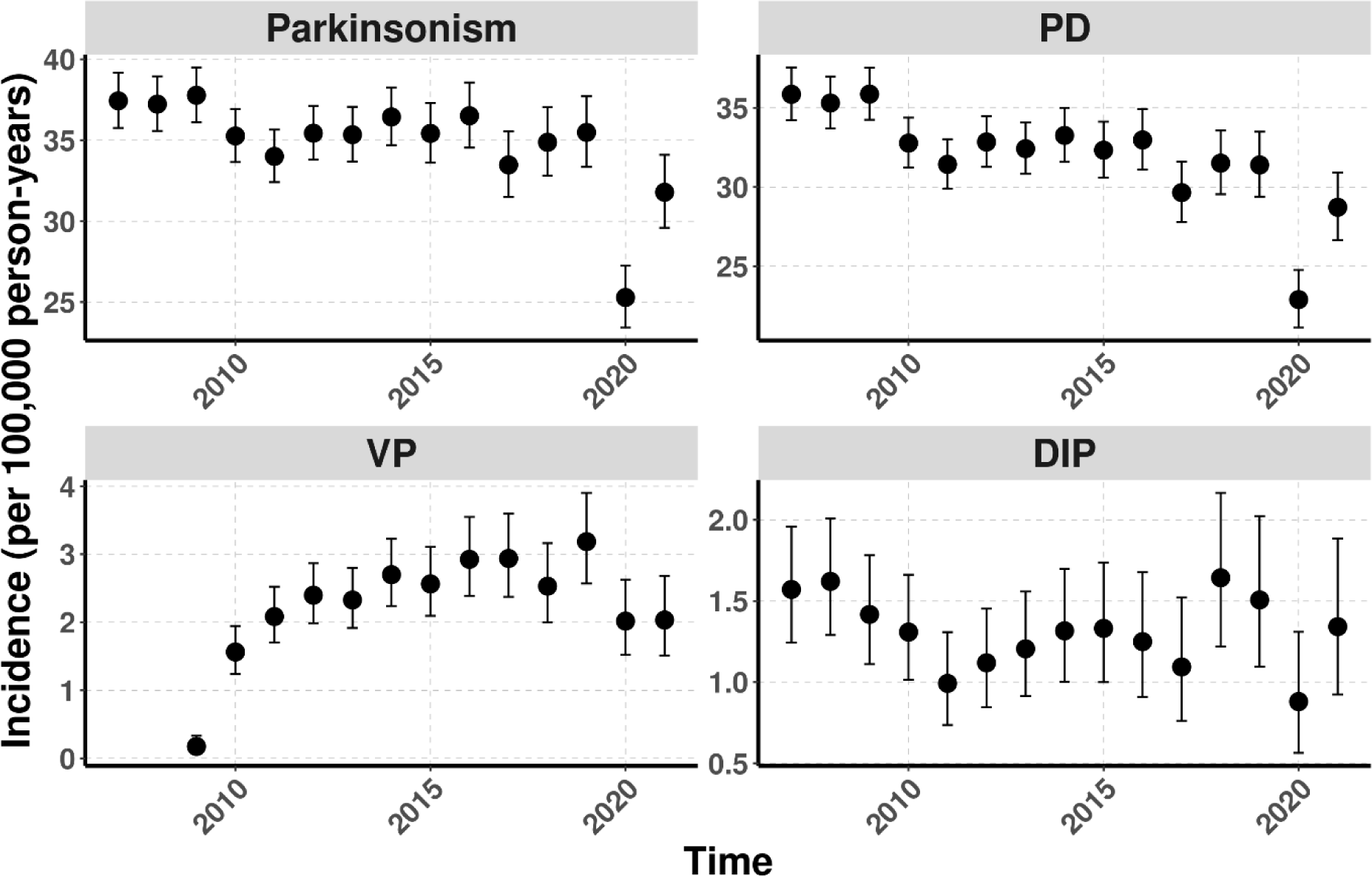
Overall annual incidence of parkinsonism, PD, VP and DIP in CPRD GOLD from 2007 to 2021

All stratified incidence results can be viewed and downloaded from an interactive Shiny web application (https://dpa-pde-oxford.shinyapps.io/ParkinsonismIncidencePrevalenceShiny/).

When stratifying by sex, incidence of parkinsonism and PD were higher in males than females (Figure 2). From 2007 to 2019, the incidence of parkinsonism and PD decreased for females whereas the incidence for males remained relatively stable. For example, the incidence of parkinsonism for females was 30.65 (28.54 – 32.88) and 27.03 (24.44 – 29.82) per 100,000 pys, respectively, in 2007 and 2019; whereas the same incidence for males was 44.43 (41.84 – 47.14) and 44.16 (40.80 – 47.74) in 2007 and 2019. The annual incidence of VP for females was comparably lower than males; the incidence of VP for females fluctuated whereas the same incidence for males an overall increase was observed. The incidence of DIP for both sexes fluctuated but the incidence for females mostly remained slightly higher than males throughout the study period.

**Figure 2:**
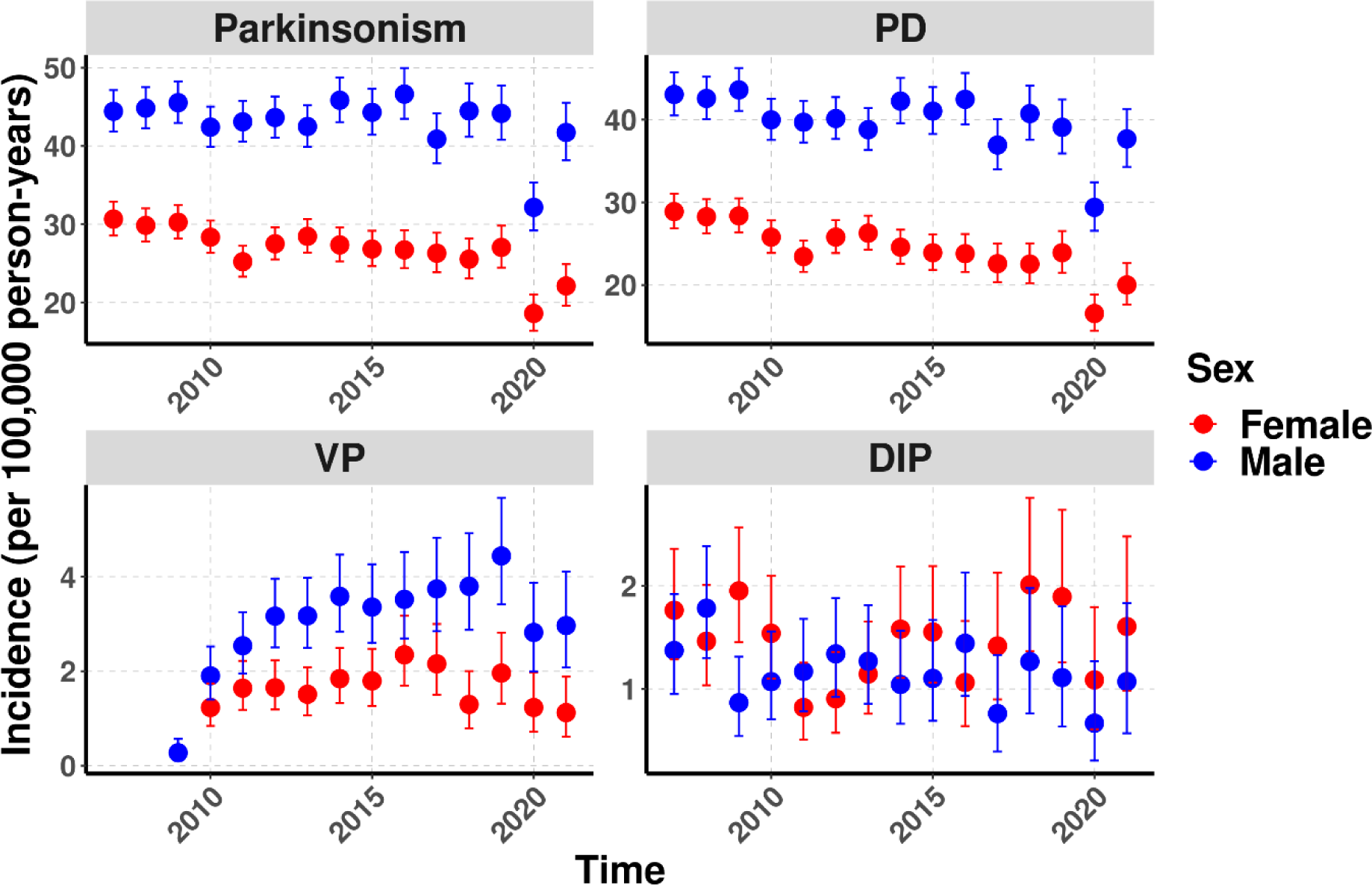
Annual incidence of parkinsonism, PD, VP and DIP stratified by sex from 2007 to 2021

When stratifying by age, the incidence of parkinsonism and subtypes increased with age (Supplementary Figures 1-4). The rates were stable for those under 70 years of age. For those aged over 70, the incidence of parkinsonism and PD decreased whereas the incidence of VP increased. For example, for those aged over 80, the incidence of PD was 207.80 per 100,000 pys (190.76 – 225.95) in 2007 and 152.21 per 100,000 pys (133.96 - 172.25) in 2019 and the incidence of VP was 13.06 (9.15 – 18.08) and 29.40 (21.75 – 38.86) in 2010 and 2019 respectively. The rates for DIP were relatively stable for all age bands considered.

When stratifying by age and sex, the incidence of parkinsonism, PD and VP remained higher for males than females across all age bands (Supplementary Figures 5-8). For both sexes aged between 71 and 80 and above 80, the incidence of parkinsonism and PD of each strata decreased whereas the incidence of VP increased from 2007 to 2019. Since the number of DIP cases were low, no trends were observed; however, the rates were often higher for females than males within the same age band.

### Crude Prevalence

From 2007, the annual prevalence of parkinsonism and PD gradually increased to reach 0.25% (0.25% - 0.26%) and 0.23% (0.23% - 0.24%) in 2016 respectively (Figure 3). Both then dipped in 2017 and 2018 before peaking in 2019 with a decline in 2020 and 2021. There was an increasing recording of VP from 2010 onwards, with prevalence being largely stable from 2017 onwards. The prevalence of DIP remained stable (0.002% to 0.003%) throughout the study period.

**Figure 3:**
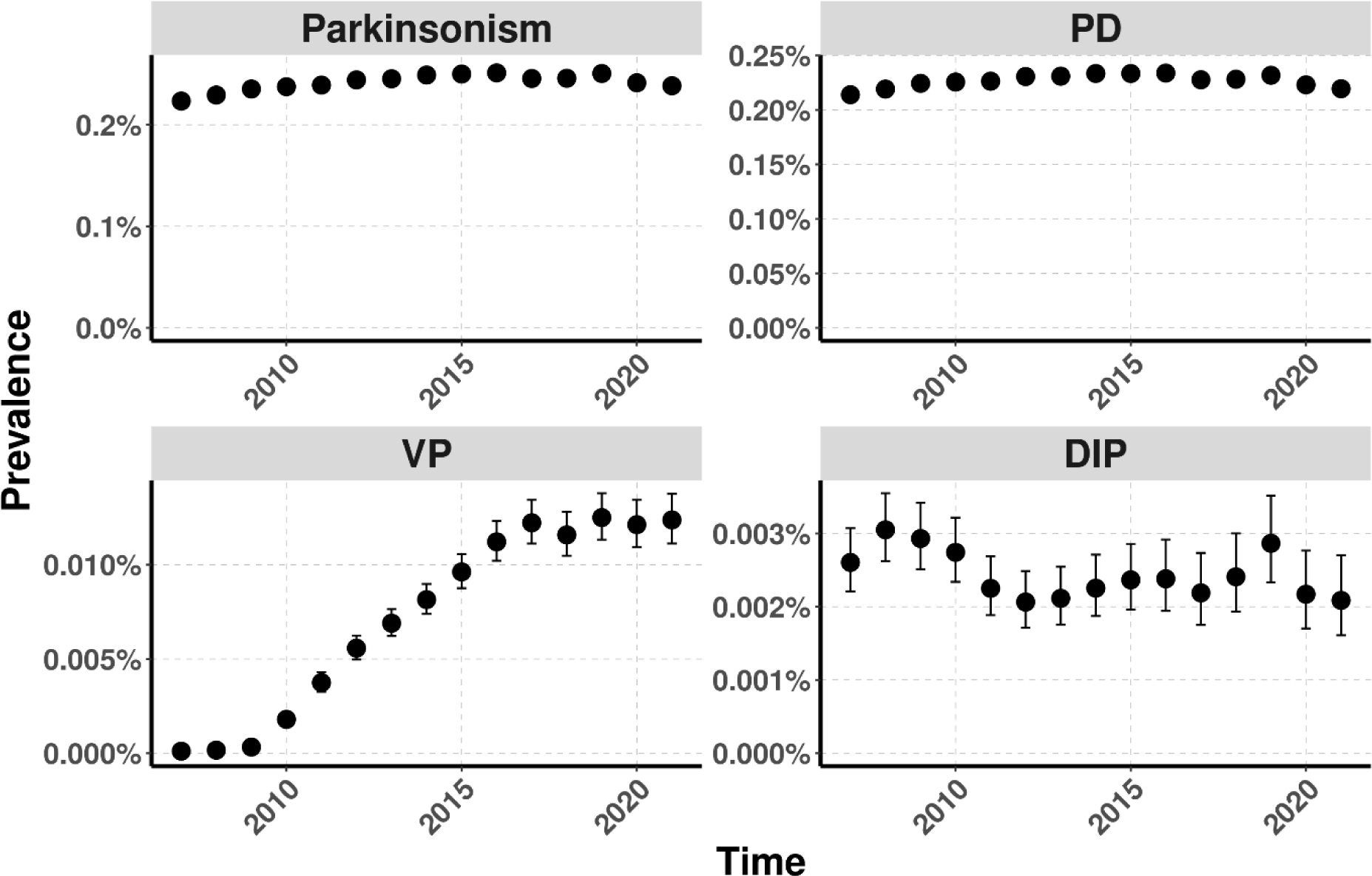
Overall annual prevalence of parkinsonism, PD, VP and DIP in CPRD GOLD from 2007 to 2021

Prevalence of parkinsonism and PD were consistently higher in males than females (Figure 4). Males showed increasing trends whereas females showed stable trends from 2007 to 2019. For example, the prevalence of PD for males was 0.25% in 2007 and increased to 0.29% in 2019 whereas the same prevalence for females was both 0.18% in 2007 and 2019. The prevalence for both sexes then declined from 2019 to 2021. For VP, the number of recordings for both sexes increased from 2010 until 2019, where both remained stable thereafter. The prevalence of DIP for both sexes remained relatively stable with females having slightly higher values than males throughout the study period; the prevalence mostly fluctuated between 0.002% to 0.004% for females and 0.001% to 0.003% for males.

**Figure 4:**
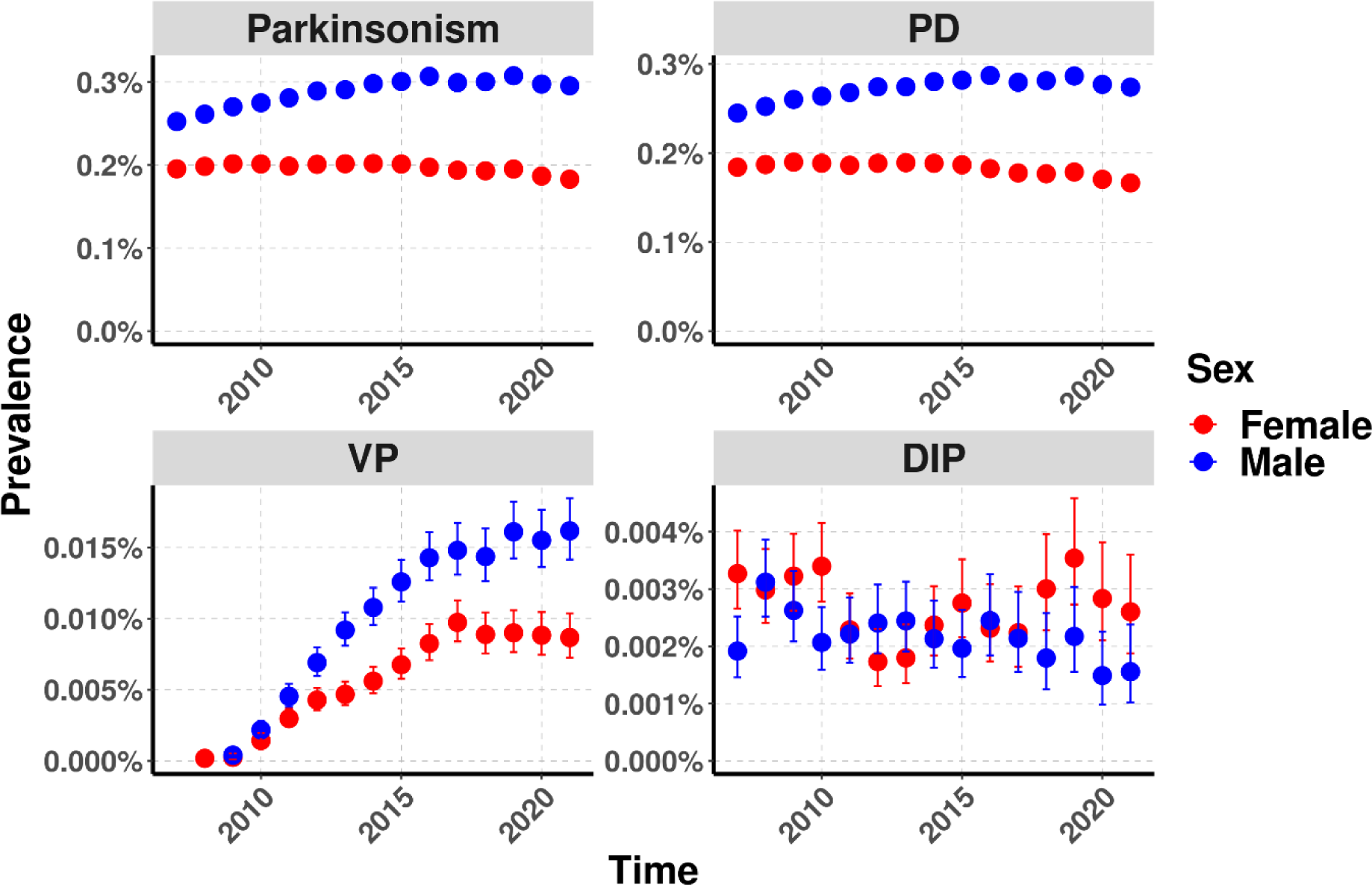
Annual prevalence of parkinsonism, PD, VP and DIP stratified by sex from 2007 to 2021

The prevalence of parkinsonism and subtypes increased with age, though such effect was not as strong for DIP (Supplementary Figures 9-12). The prevalence of parkinsonism and PD was either stable or it increased slightly and then decreased slightly. For example, for the 71-80 age band, the prevalence of parkinsonism and PD increased from 2007 (parkinsonism: 0.99%, PD: 0.96%) to 2012 (parkinsonism: 1.08%, PD: 1.03%) and decreased from 2012 to 2021 (parkinsonism: 0.93%, PD: 0.87%). The prevalence of VP remained stable for people under 70-years of age whereas the same prevalence for people aged over 70 increased steadily throughout the study period. For example, the prevalence for the 80+ age band started with 0.01% in 2010 (data was insufficient in 2007 to 2009), it increased to 0.11% in 2021 and peaked at 0.12% in 2019. The prevalence of DIP for each age band remained stable.

When stratifying by both age and sex, the prevalence of parkinsonism and PD showed an overall decreasing trend for females aged between 61 and 70 and an overall increasing trend for males in the same age band. The annual prevalence of PD for females aged between 71 and 80 and both males and females aged above 80 all showed an overall decrease. For people over 80-years of age, the prevalence of VP in males showed a larger increase over time compared to females. The prevalence of DIP remained stable across all strata (Supplementary Figures 13-16).

### Age-standardized Incidence and Prevalence

Age-standardized incidence and prevalence of parkinsonism and subtypes followed the same secular trends as the crude results, for the whole population and when stratified by sex (Supplementary Tables S3-S4, Supplementary Figures 17-20).

## Discussion

### Key results

This study provides a comprehensive assessment on the secular changes of the incidence and prevalence and characterization of those with a diagnosis of parkinsonism, PD, VP and DIP in the UK. The incidence of parkinsonism and PD have slightly decreased over time with the prevalence slightly increasing. The number of VP diagnoses increased progressively from 2010 to 2018 and remained stable thereafter. The incidence and prevalence of DIP remained stable over time. The incidence and prevalence of PD and VP were higher in males than females and such disparity increased with age.

### Demographic differences among individuals with parkinsonism subtypes

In line with previous studies, the most frequently observed parkinsonism subtype was PD [11,12,20]. The second most common subtype was VP, contributing ∼5% of total parkinsonism cases, echoing The Rotterdam Study [20]. DIP contributed the least to total parkinsonism cases in this work, which contrasts with some previous studies that state DIP is the second most prevalent subtype of parkinsonism [5,21]. This could be explained by the underdiagnosis of DIP as a medical condition [6] and misclassification of DIP as PD [5]. Also, in line with literature, this study shows that males were twice as likely to develop PD and VP than females [22–25], whereas DIP was slightly more common in females [6,21,26], and that the median age of onset of VP was later than PD [27,28]. However, there are conflicting results about the average age of DIP onset compared to PD; this work and other studies have shown lower average age of DIP onset compared to PD [29–31] whilst another study has showed the opposite [5]. Such contrast could be attributed partially to the large variation in age among people who use these causative medications. For example, although antipsychotics are used more frequently in older populations [32], studies have shown that there has been an increasing trend in antipsychotics prescriptions in children and adolescents [33,34].

Since VP is characterized by having a previous episode of ischemic cerebrovascular disease before showing Parkinson-like symptoms, it is unsurprising that these patients have a higher proportion of cardiovascular diseases compared to patients diagnosed with PD or DIP due to the overlapping risk factors and etiologies between cardiovascular diseases and cerebrovascular diseases [35,36]. Although it is also unsurprising that the number of stroke cases is higher in VP patients compared to other subtypes, the percentage of VP patients diagnosed with stroke any time prior to VP diagnosis was low (<20%). This could be potentially explained by under-recording of stroke in primary healthcare settings [37]. Works have also demonstrated an association between cardiovascular disease and an increased risk of dementia [38], and that patients with CKD are more at risk of stroke [39], this may explain why the number of dementia and CKD cases are higher among VP patients.

Regarding DIP, these patients showed higher proportions of medications such as psycholeptics, antileptics and antidepressants one year before DIP diagnosis. These medications are known dopamine receptor antagonists with strong evidence of inducing Parkinson-like symptoms [5,6].

### Incidence and prevalence of parkinsonism and PD

Since PD cases contributed the most to total parkinsonism cases, the temporal trends of PD largely mirror the trends of parkinsonism.

Prior studies in the UK reported much higher incidence values than our study [10,40]. For example, Okunoye *et al*. reported an incidence rate of 64.8 per 100,000 pys for PD in 2015 [10], whilst the same incidence rate was 32.3 per 100,000 pys in our study. The reason for this difference is due to the different age distribution of the study population. Our study includes those aged 18 and older with Okunoye *et al*. including those aged 50 and older. For our study, the age-specific crude incidence rate for those aged above 50 ranged from ∼15 per 100,000 pys in those aged between 51 to 60 to ∼160 per 100,000 pys in those aged above 80. On the other hand, in line with previous literature, our findings on secular trends show that the annual incidence of PD has decreased in the UK over time [10,40], and one potential reason for this slight decrease is the change of PD recordings in primary healthcare settings [40]. Regarding prevalence, our results are in line with a Reference Report from Parkinson’s UK [41], also using CPRD GOLD, which estimated the prevalence of PD in 2015 at 0.27% in people aged over 18 compared to a prevalence of 0.23% from our work. Aligning with other studies [42,43], our work shows that the prevalence of PD has increased over time and a potential drive for this increase might be better treatment and survival [43].

Concordant with existing literature, incidence and prevalence of PD are found to be higher in males [10,30,40,44]. Factors that may have contributed to such difference include genetics [45], a greater chance of developing traumatic head injury [46], and a greater exposure to adverse environmental factors due to working in industry sectors [47]. As expected, stratifying by age shows that incidence and prevalence of PD are higher in older age groups, and this can be explained by the nature of PD, which is characterized by the gradual and progressive presence of Lewy Bodies and loss of dopamine-producing neurons in the brain over time [2].

The incidence and prevalence of PD both decreased in 2020 and 2021 and a key factor behind this decline is the reduction of diagnosis during the pandemic. Delayed diagnoses will result in poorer outcomes, potentially increasing the number of Parkinson’s disease-related deaths during the pandemic [48], and this also plays a role in reducing the prevalence of PD.

### Incidence and prevalence of VP

The increase of incidence and prevalence of VP could be attributed to a better recognition of this atypical disorder in the UK [40]. As a medical condition, VP has been heavily debated over the years [27,49]. It has been established as a new entity only in 2001 [50], and the first set of strict clinical criteria of VP been only established in 2004 [51]. Our study shows that the incidence and prevalence of VP are higher in males and one reason that could explain this finding is higher incidence of stroke in males [52], as stroke often precedes VP [27]. Moreover, it has also been shown that the severity of strokes tends to be higher in females, who are typically older and have higher mortality rates [53]. This may further reduce incidence and prevalence of VP for females, older age of stroke onset and higher mortality from stroke in females may mean a lesser chance of developing VP. This disparity between sexes increased with age, and it could be explained by males being more likely to experience multiple strokes [54] and that the incidence of recurrent stroke increases with age [29].

### Incidence and prevalence of DIP

In contrast to PD and VP, this study finds that DIP has a higher incidence and prevalence among females, which agrees with previous literature [5,31,55]. Reasons for this are multifaceted and likely to involve a combination of biological, pharmacokinetic, and social/behavioral factors. Females typically have higher body fat and lower body water percentages, which affects drug distribution and metabolism as well as hormonal differences such as estrogen that has been shown to influence drug metabolism [56,57]. Furthermore, females are also more likely to be prescribed medications that can cause DIP such as antidepressants [58] and antipsychotics [32,59] and are more likely to have polypharmacy leading to risk of side effects such as parkinsonism [57]. In contrast to Han *et al*. [55] where the incidence of DIP was decreasing, incidence and prevalence of DIP were both stable in this study. There are several reasons that can explain this difference including definition of DIP (the definition in Han *et al*. included the use of causative drugs) and population characteristics (the study in Han *et al*. was based in Asia).

### Strengths and limitations

This study has many strengths. Firstly, this study used CPRD GOLD, a large primary health care database covering GPs from England, Wales, Scotland and Northern Ireland, and thus may improve generalizability across the UK. However, one may need to consider the change of the size of the database over time when interpreting the results. For example, the number of people in the database gradually decreased after 2011, likely due to GP practices in England moving to another clinical information system not captured in CPRD GOLD. Another strength of this study is that we have analyzed the trends of the incidence and prevalence in each age and sex strata. In contrast to other studies [11,12,25], the inclusion of these stratified results can be useful in better informing which groups of people require more attention in terms of prevention.

One limitation of this study could be the misclassification of the subtypes, as previous literature suggests that VP and DIP cases could be misclassified as PD by healthcare professionals [5,6,27]. This means that the actual incidence and prevalence of PD might be overestimated whilst the incidence and prevalence of DIP and VP might be underestimated. Future works exploring stricter definitions of these subtypes are strongly advised. For example, the inclusion of causative drugs could be considered when defining DIP [55]. Similarly, although the UK Parkinson’s Disease Society Brain Bank criteria has been validated to have high specificity and sensitivity [60], the addition of prescriptions of antiparkinsonian drugs such as levodopa can provide a stricter definition of PD.

## Conclusion

Though the incidence of parkinsonism and PD have been slowly decreasing, the prevalence of both have been slowly increasing, potentially suggesting improved survival and care. Both incidence and prevalence of VP have been steadily increasing which may suggest a change of diagnosis of VP and potentially a call for screening program for people who experience an episode of ischemic cerebrovascular disease. The incidence and prevalence of DIP have been stable throughout the study period. As the UK is known to have an ageing population, parkinsonism and subtypes pose an ever-increasing burden to the UK.

## Funding agency

The author(s) declare that financial support was received for the research, authorship, and/or publication of this work. This research was partially funded by the European Health Data and Evidence Network (EHDEN). This activity under the EHDEN has received funding from the Innovative Medicines Initiative 2 (IMI2) Joint Undertaking under grant agreement No 806968. IMI2 receives support from the European Union’s Horizon 2020 research and innovation program and European Federation of Pharmaceutical Industries and Associations (EFPIA). The sponsors of the study did not have any involvement in the writing of the manuscript or the decision to submit it for publication. Additionally, there was partial support from the Oxford NIHR Biomedical Research Centre.

## Abbreviations

CDM: Common Data Model
CI: Confidence interval
CKD: Chronic Kidney Disease
CPRD: Clinical Practice Research Datalink
DIP: Drug-induced Parkinsonism
IQR: Inter-quartile range
OMOP: Observational Medical Outcomes Partnership
PD: Parkinson’s Disease
pys: person-years
RAS: Renin-angiotensin-system
RDG: Research Data Governance
VP: Vascular Parkinsonism

## Data Availability Statement

This study is based in part on data from the Clinical Practice Research Datalink (CPRD) obtained under license from the UK Medicines and Healthcare Products Regulatory Agency. The data is provided by patients and collected by the NHS as part of their care and support. The interpretation and conclusions obtained in this study are those of the authors alone. Patient level data used in this study was obtained through an approved application to the CPRD (Protocol number: 22_002351) and is only available following an approval process to safeguard the confidentiality of patient data. Details on how to apply for data access can be found at https://cprd.com/data-access.

## Acknowledgements

None.

## Authors’ Roles

All authors were involved in the study conception and design, interpretation of the results, and the preparation of the manuscript. AD and WYM implemented data curation, data quality tests and assessment. All authors had access to the CPRD data. XC carried out data analysis for the manuscript. DPA and LMP reviewed the clinical code lists for this study. XC and DN wrote the initial draft of the manuscript, and all authors critically reviewed the final manuscript and gave consent for publication.

