## supplementary_figures for "Secular trends of incidence and prevalence of parkinsonism and subtypes: A cohort study in the United Kingdom"

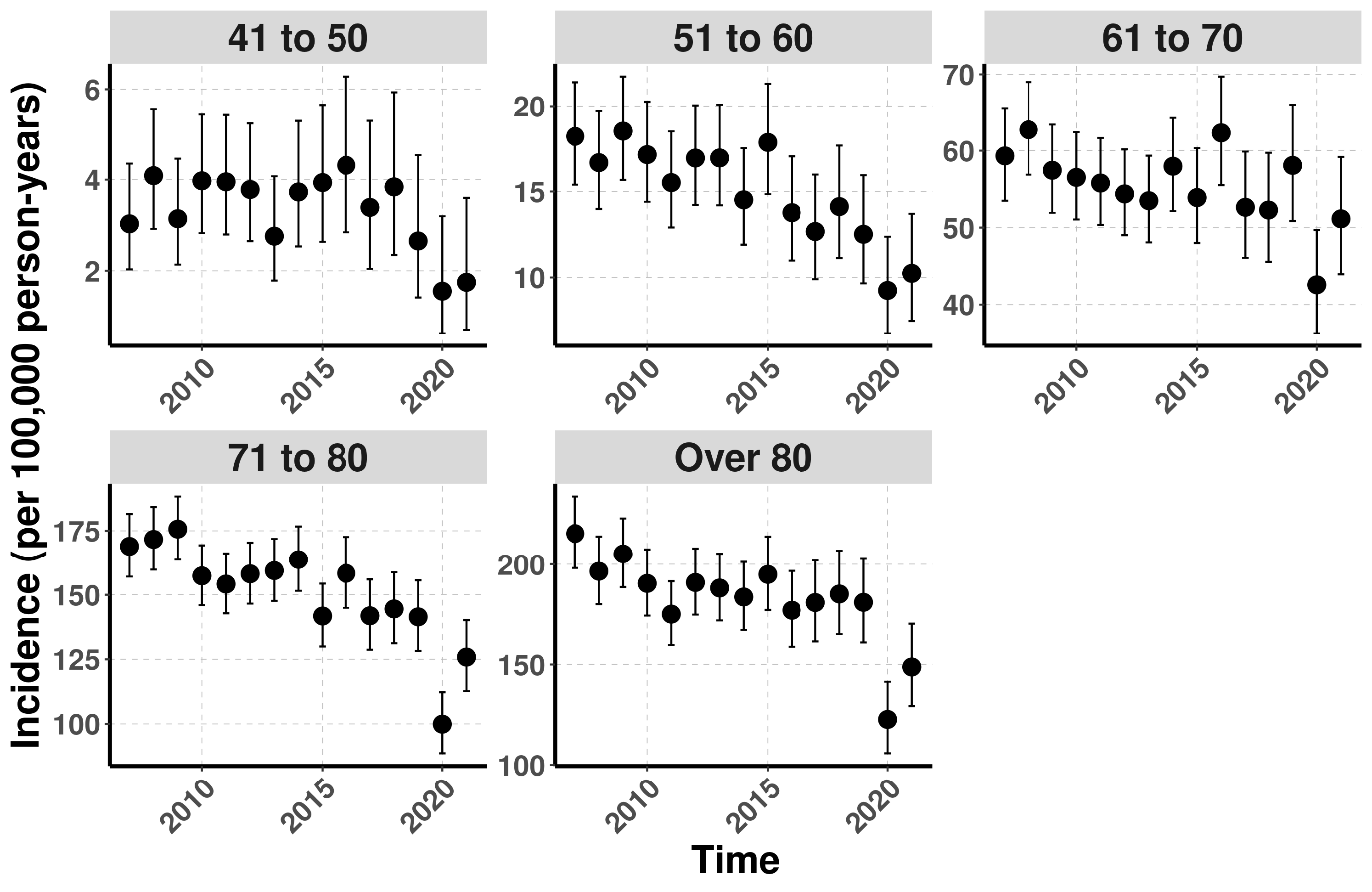

**Supplementary Figure 1: Crude annual incidence of parkinsonism stratified by age from 2007 to 2021**

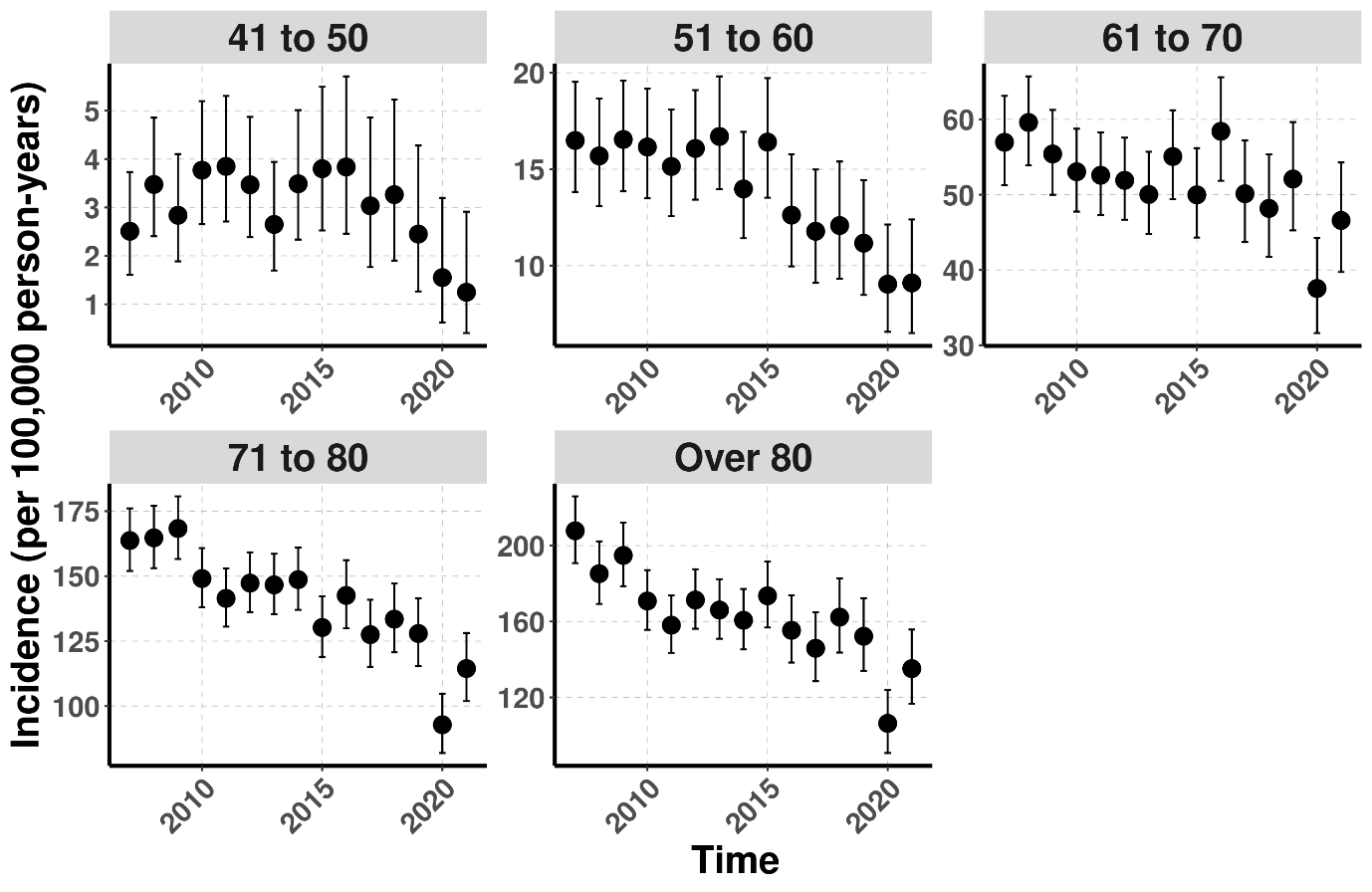

**Supplementary Figure 2: Crude annual incidence of PD stratified by age from 2007 to 2021**

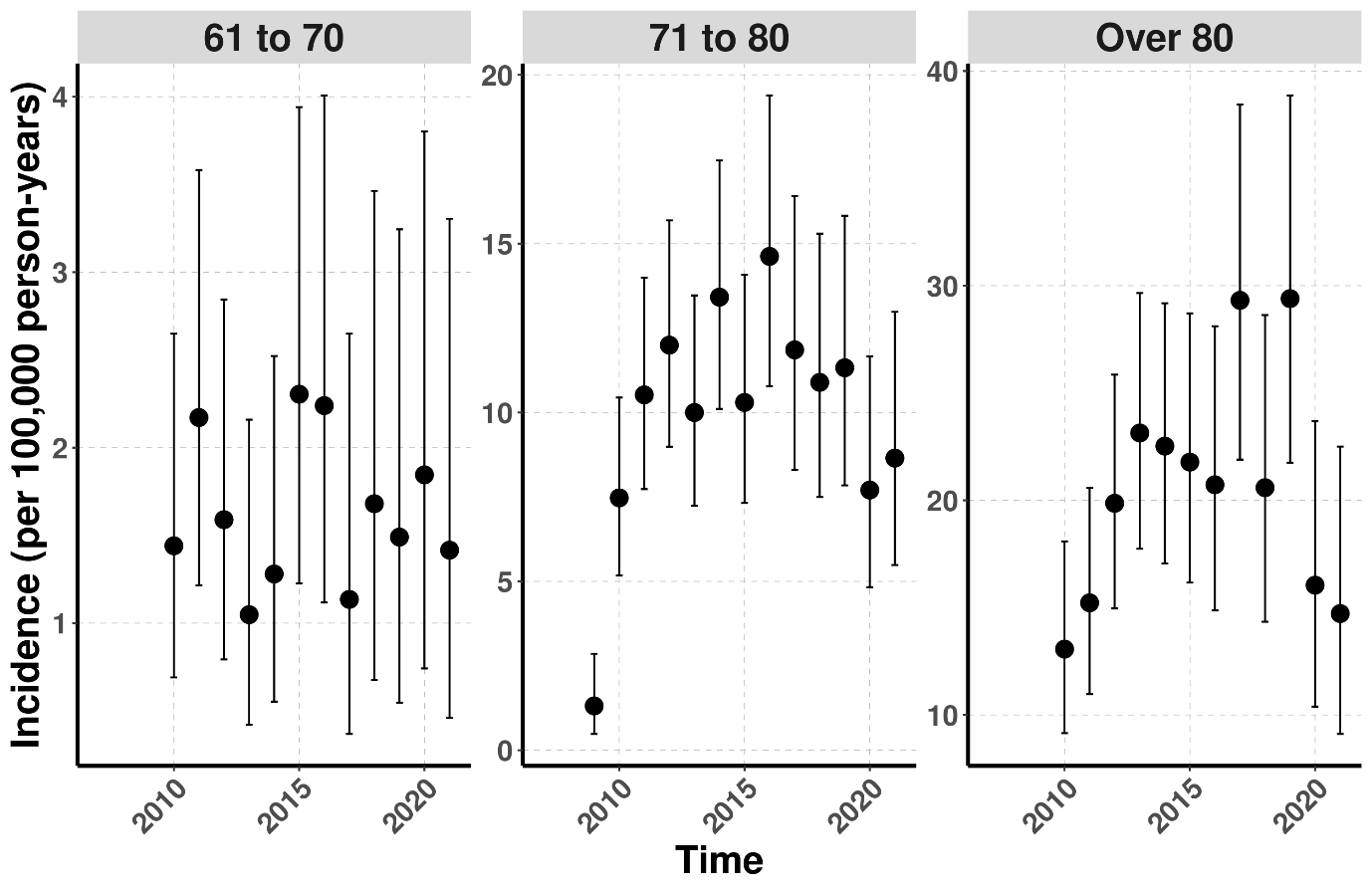
**Supplementary Figure 3: Crude annual incidence of VP stratified by age from 2007 to 2021**

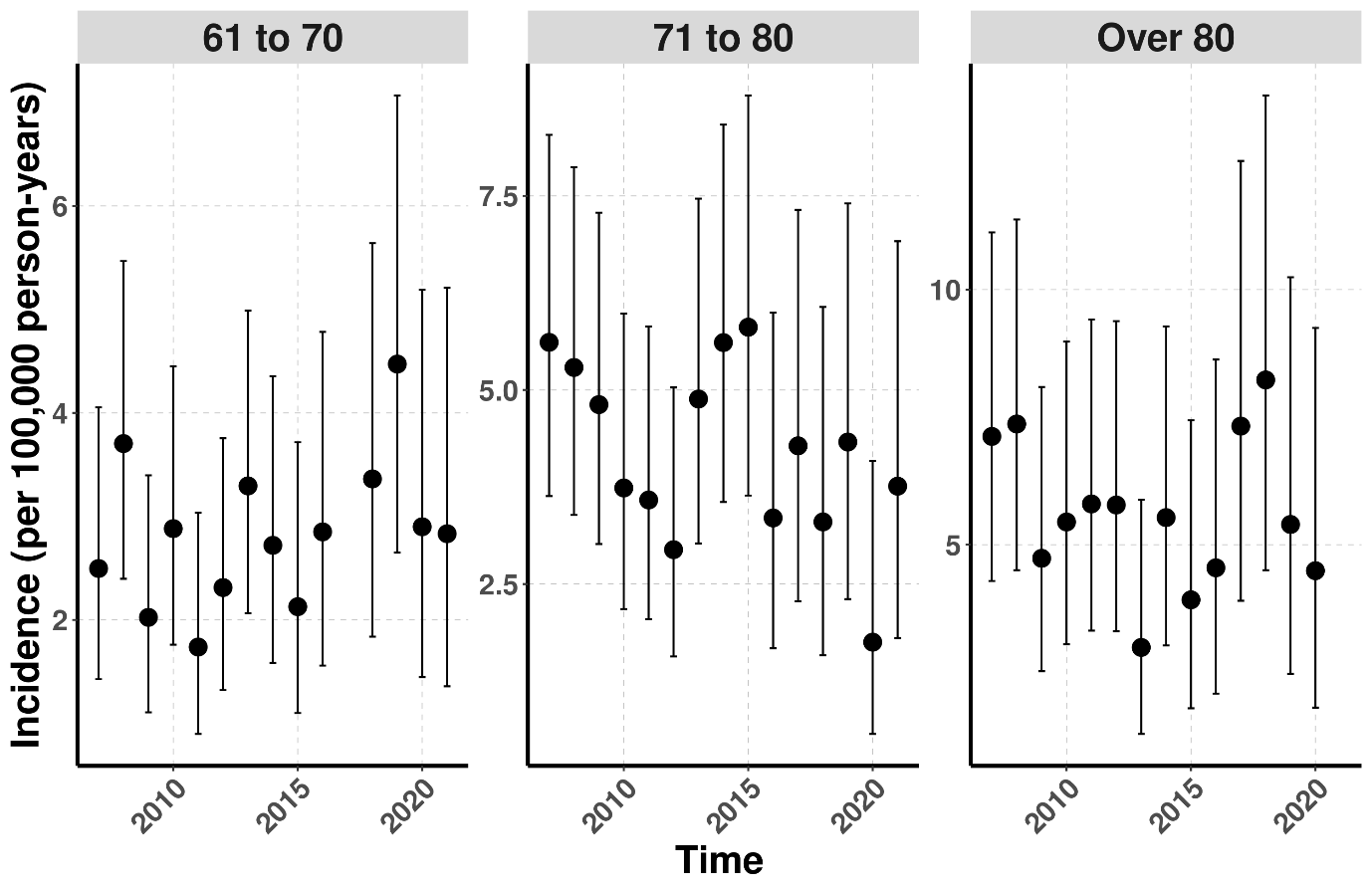

**Supplementary Figure 4: Crude annual incidence of DIP stratified by age from 2007 to 2021**

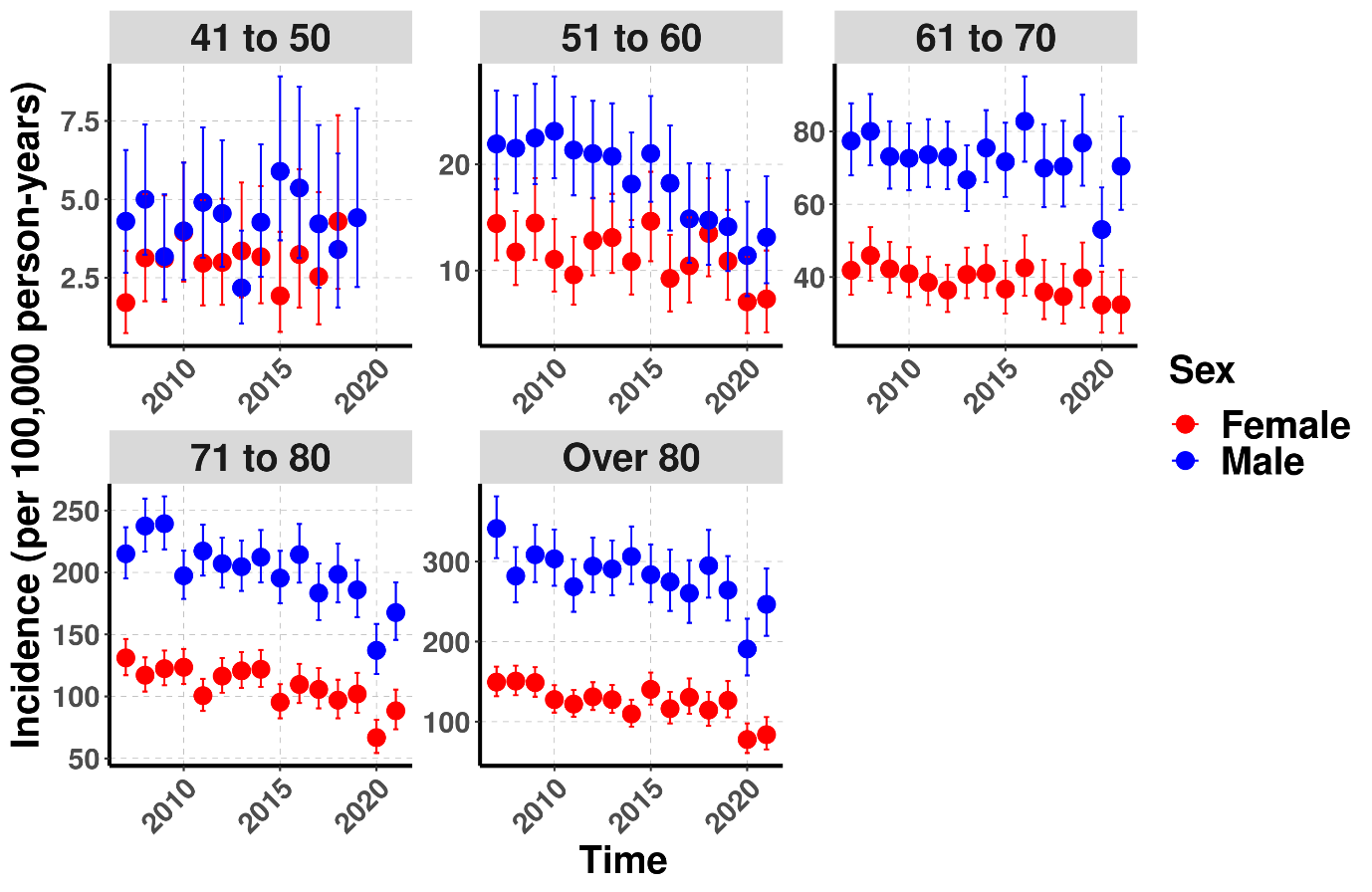

**Supplementary Figure 5: Crude annual incidence of parkinsonism stratified by age and sex from 2007 to 2021**

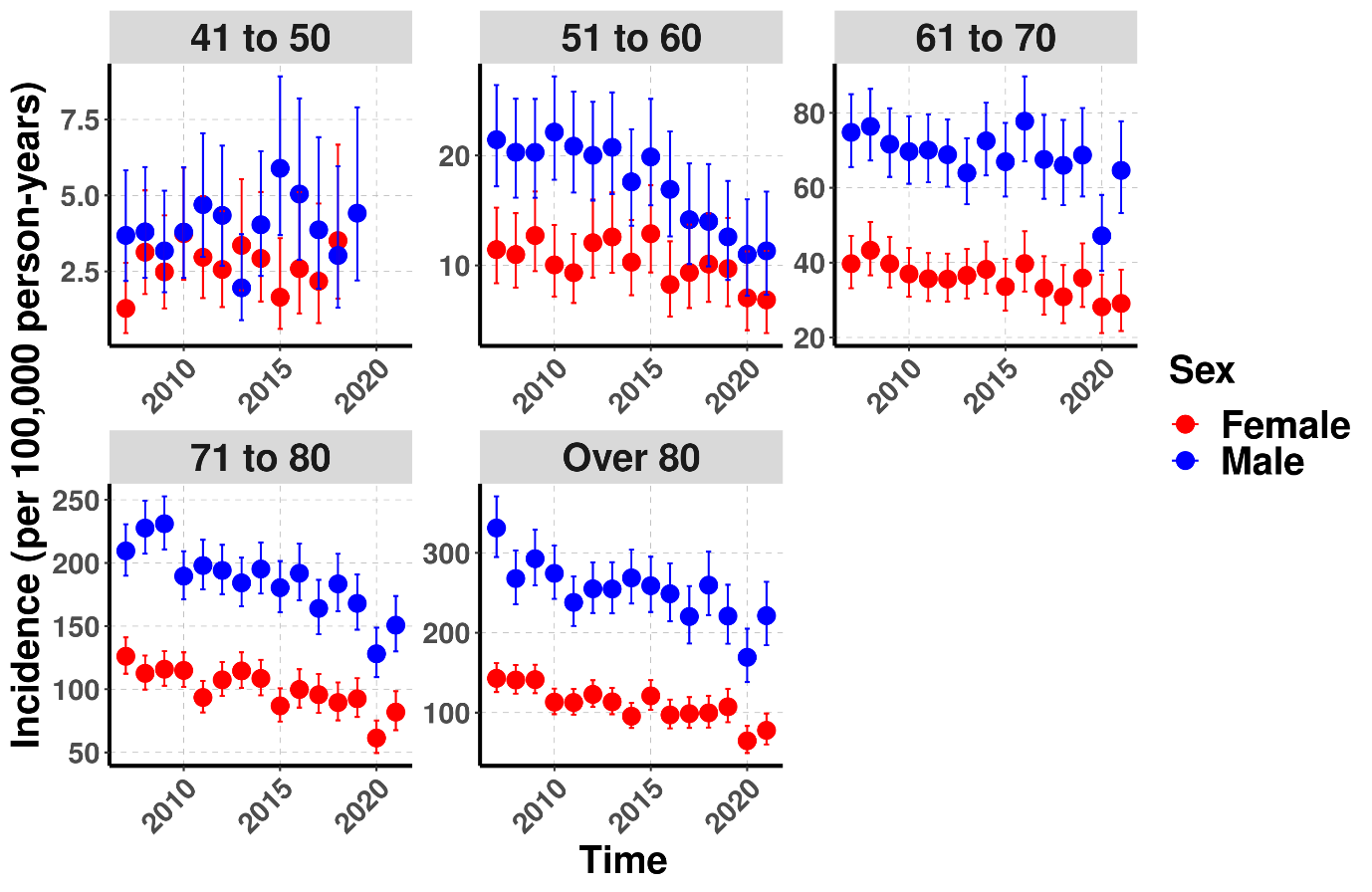

**Supplementary Figure 6: Crude annual incidence of PD stratified by age and sex from 2007 to 2021**

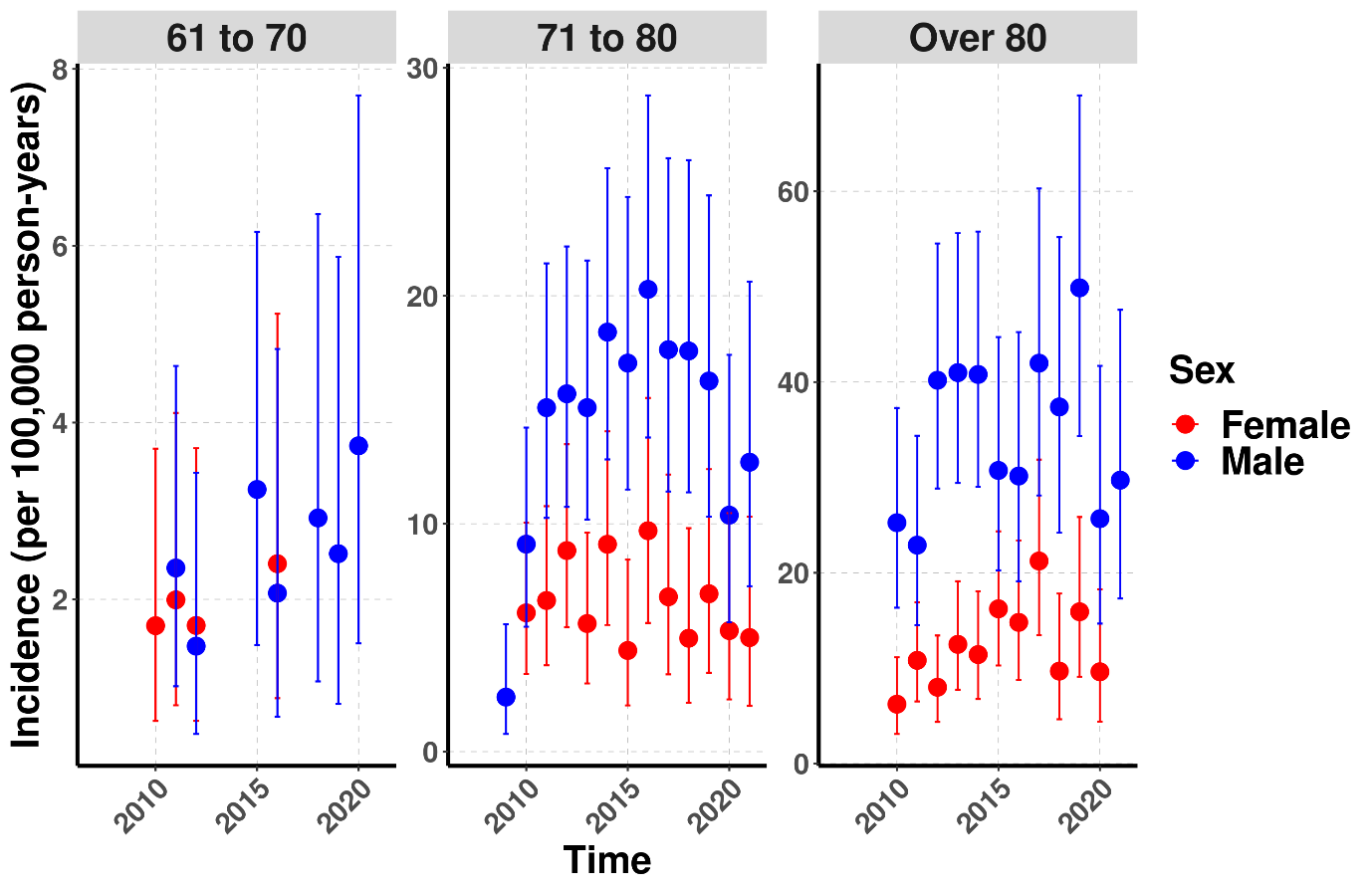

**Supplementary Figure 7: Crude annual incidence of VP stratified by age and sex from 2007 to 2021**

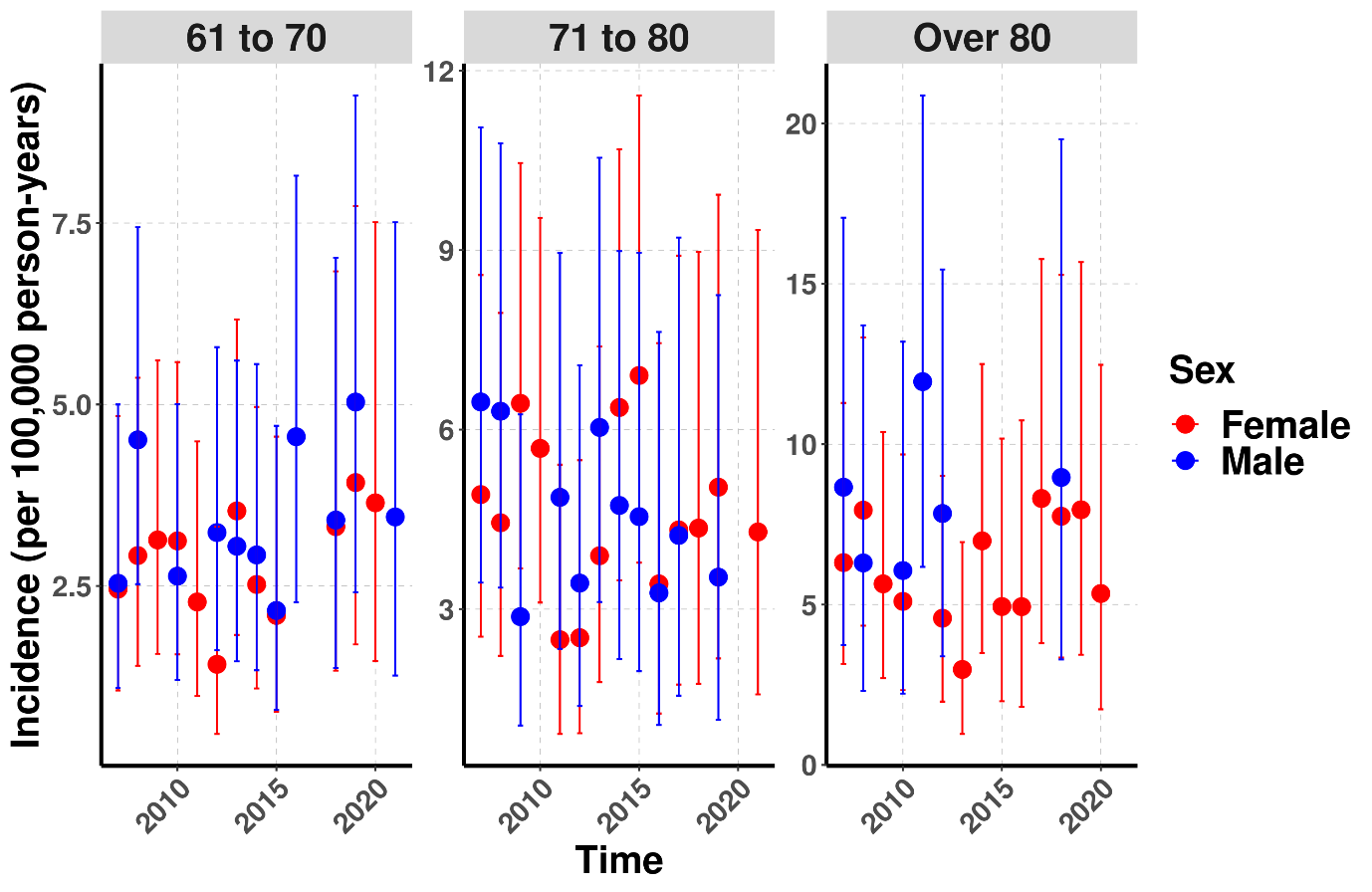

**Supplementary Figure 8: Crude annual incidence of DIP stratified by age and sex from 2007 to 2021**

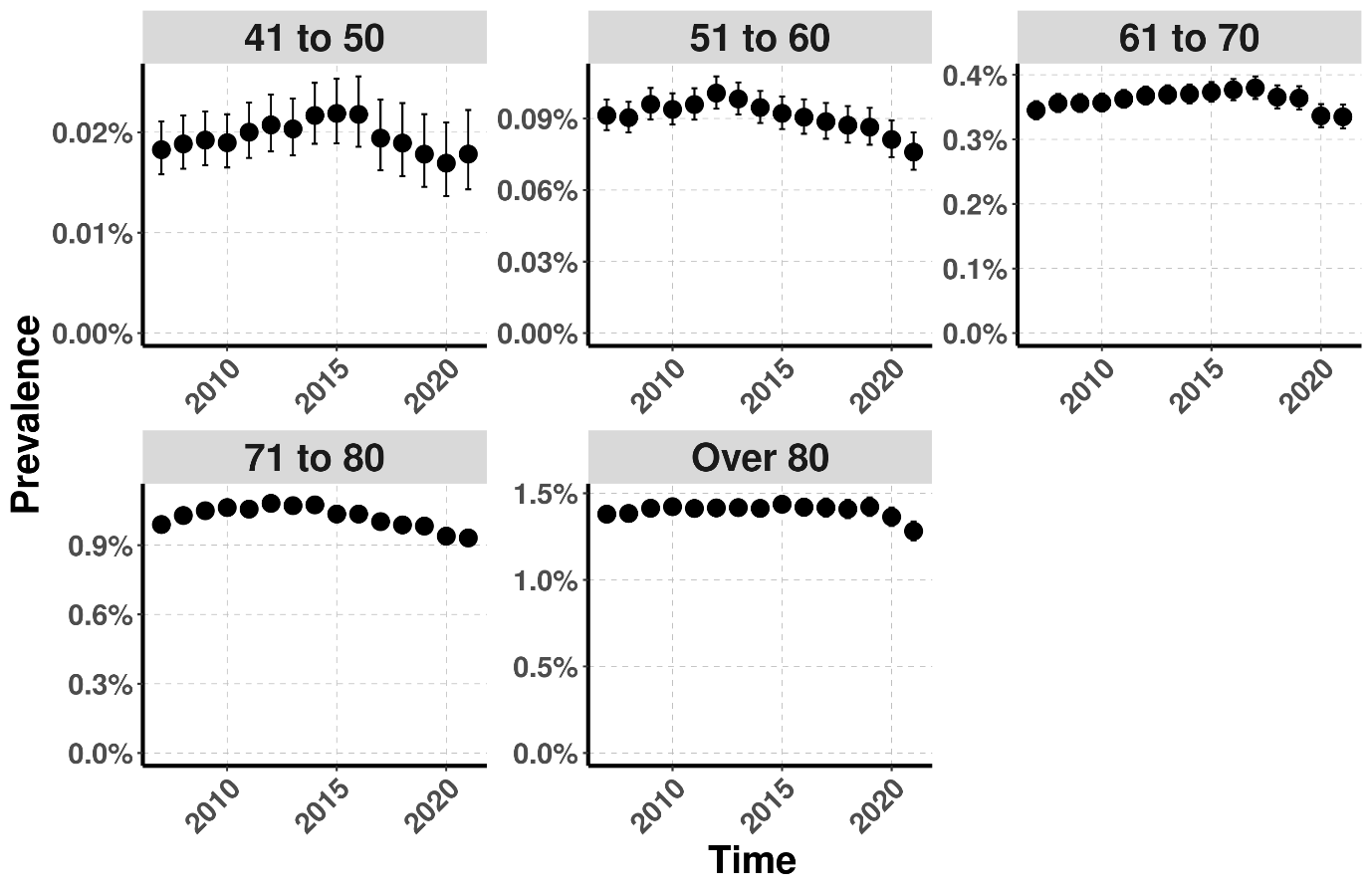

**Supplementary Figure 9: Annual prevalence of parkinsonism stratified by age from 2007 to 2021**

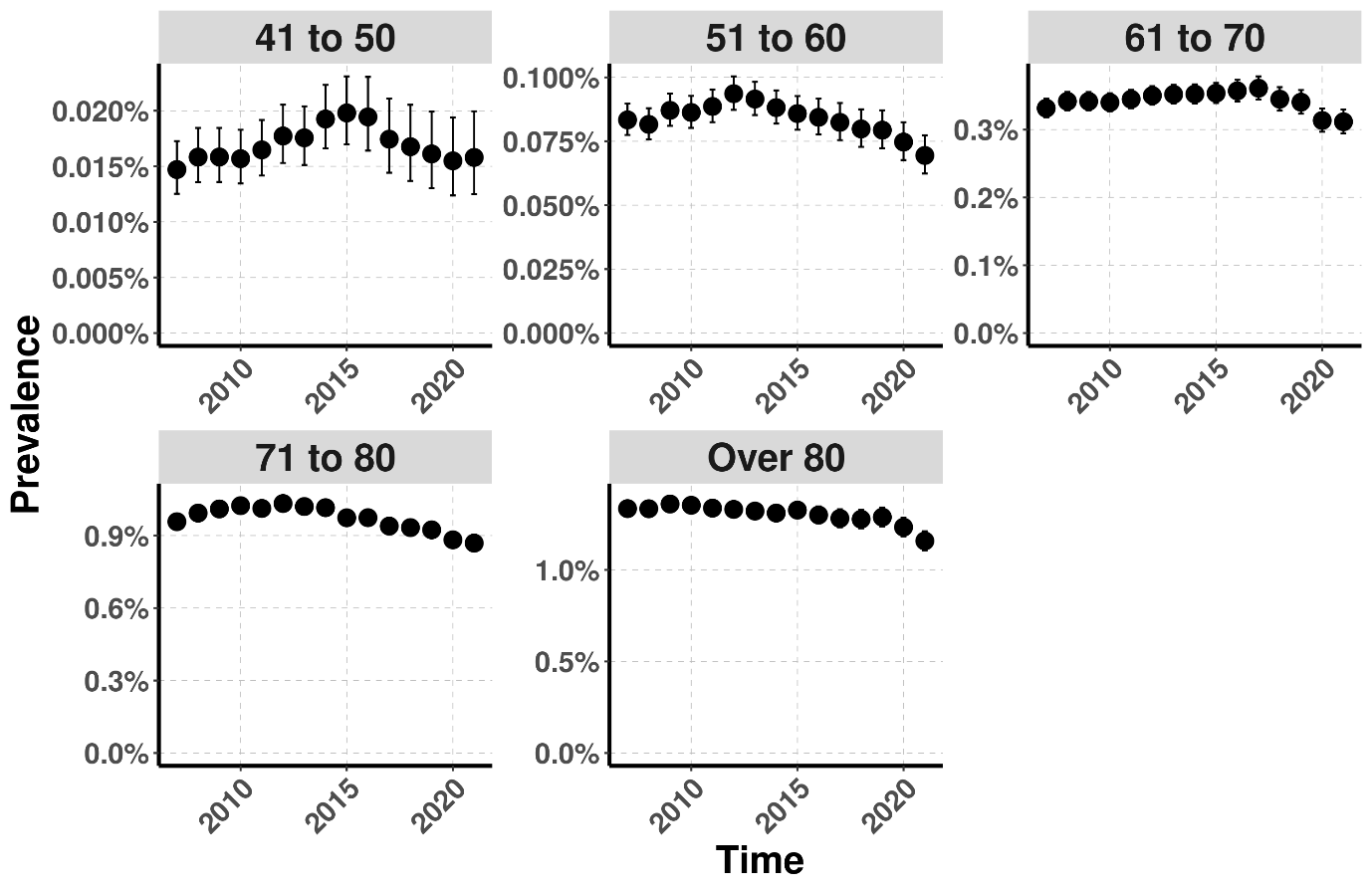

**Supplementary Figure 10: Annual prevalence of PD stratified by age from 2007 to 2021**

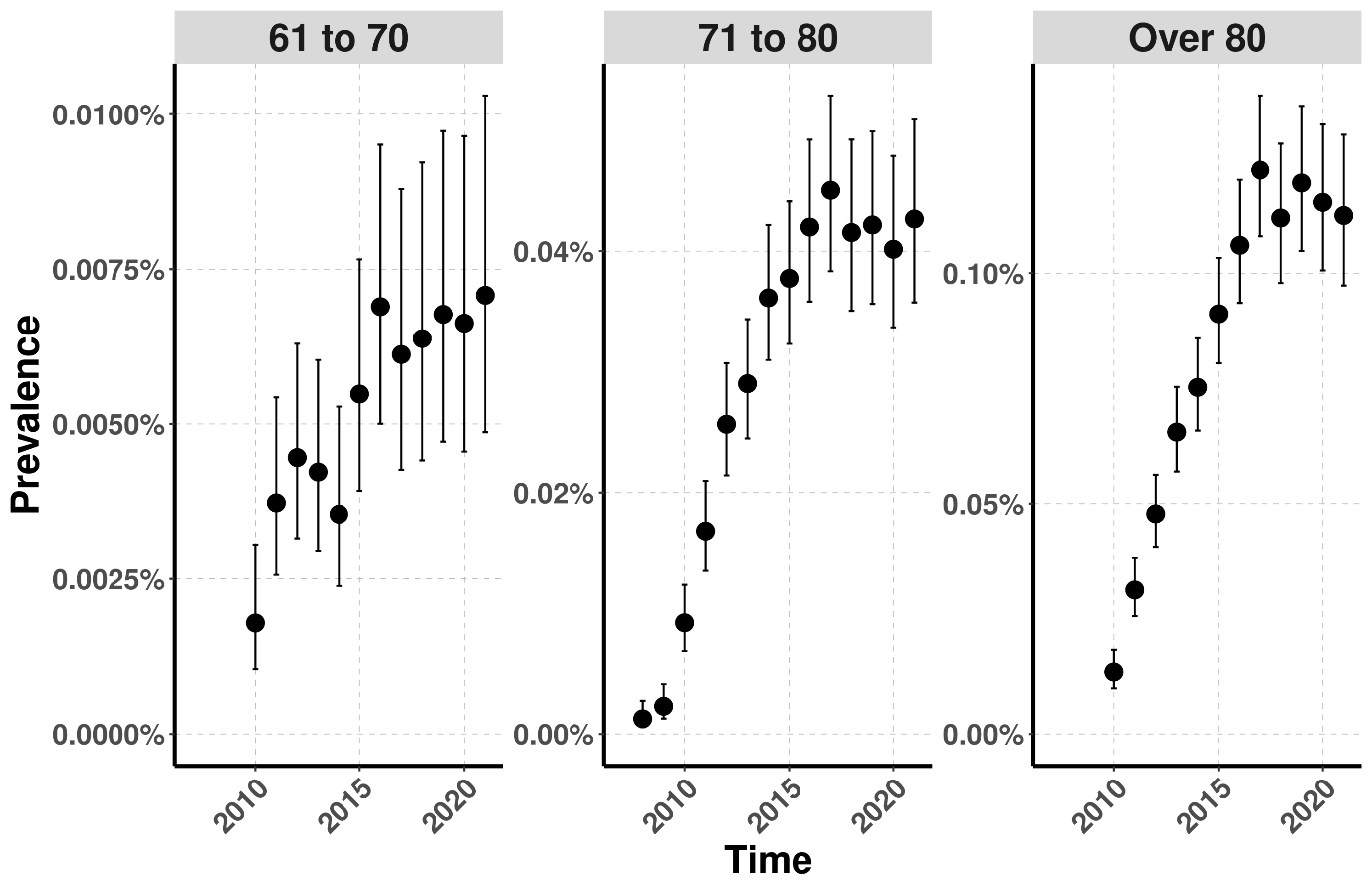

**Supplementary Figure 11: Annual prevalence of VP stratified by age from 2007 to 2021**

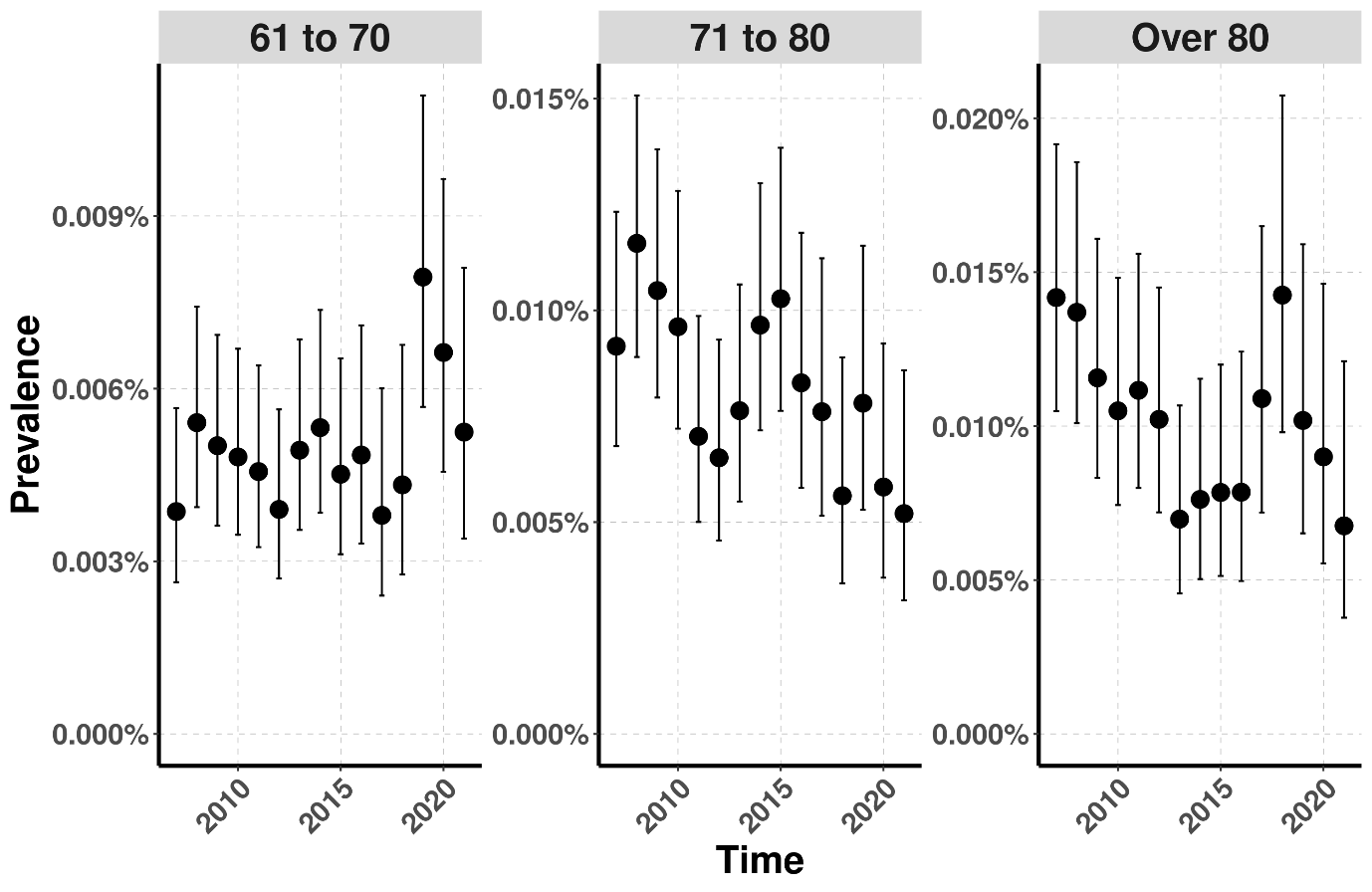

**Supplementary Figure 12: Annual prevalence of DIP stratified by age from 2007 to 2021**

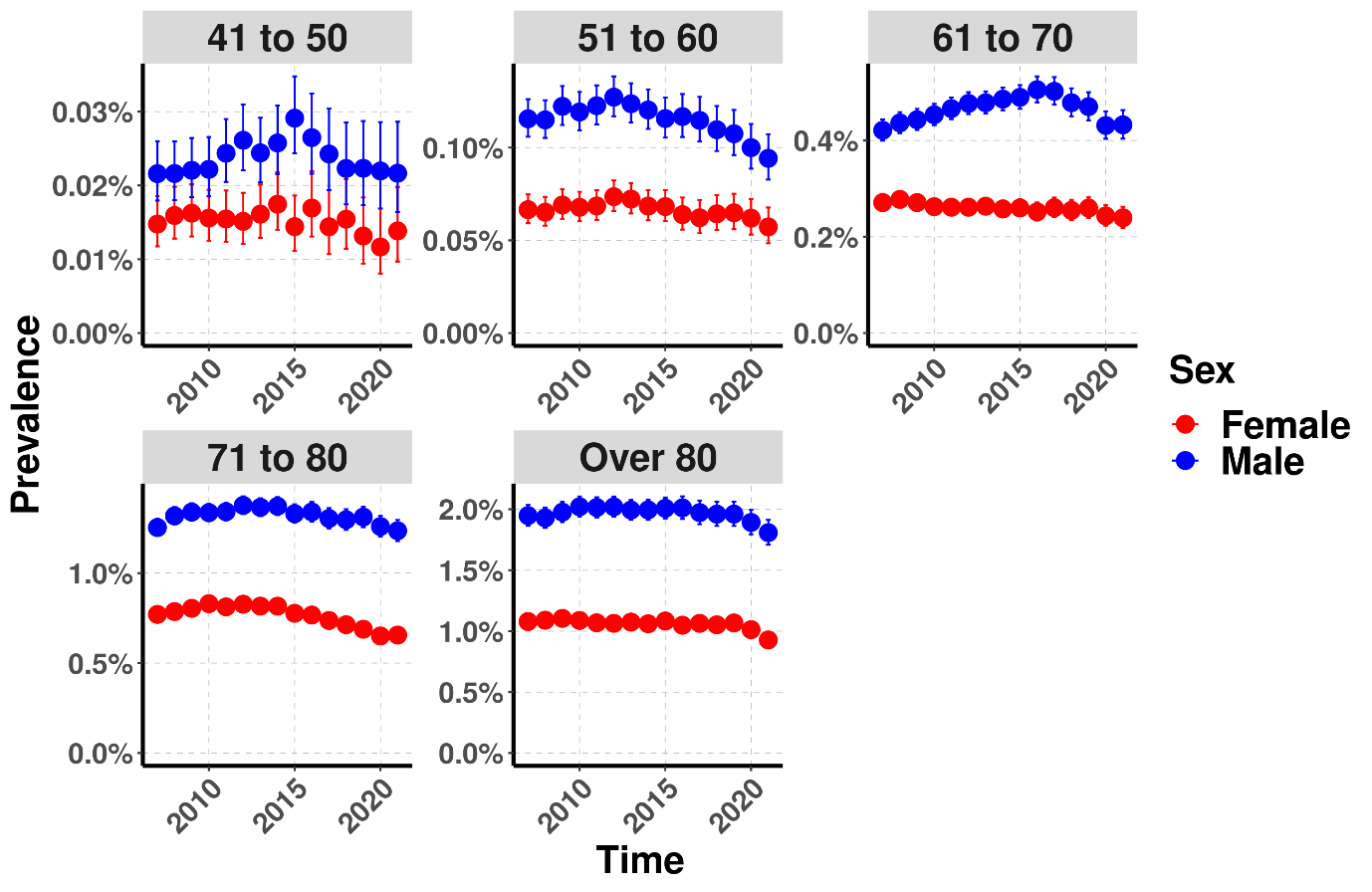

**Supplementary Figure 13: Prevalence of parkinsonism stratified by age and sex from 2007 to 2021**

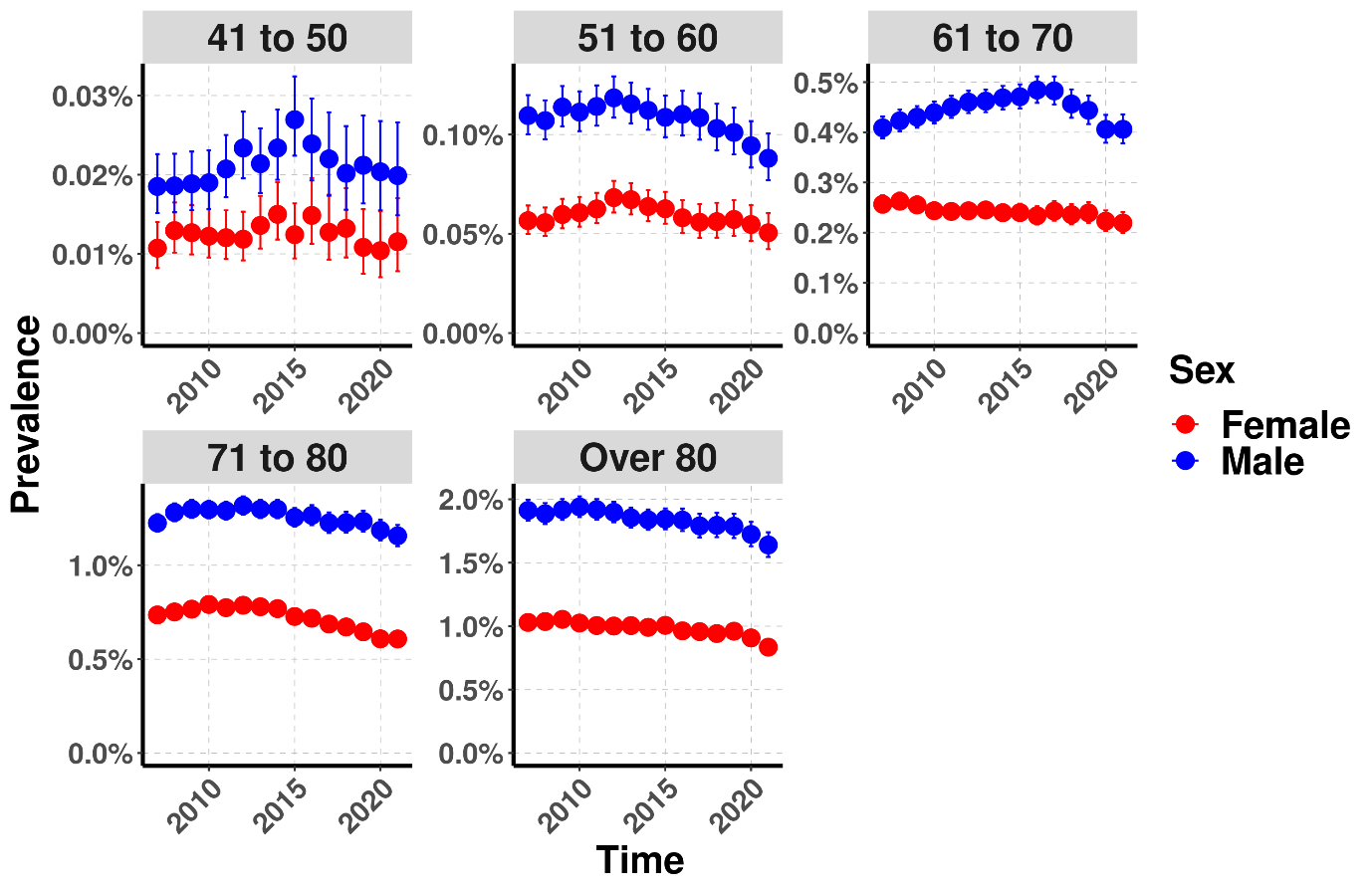

**Supplementary Figure 14: Annual prevalence of PD stratified by age and sex from 2007 to 2021**

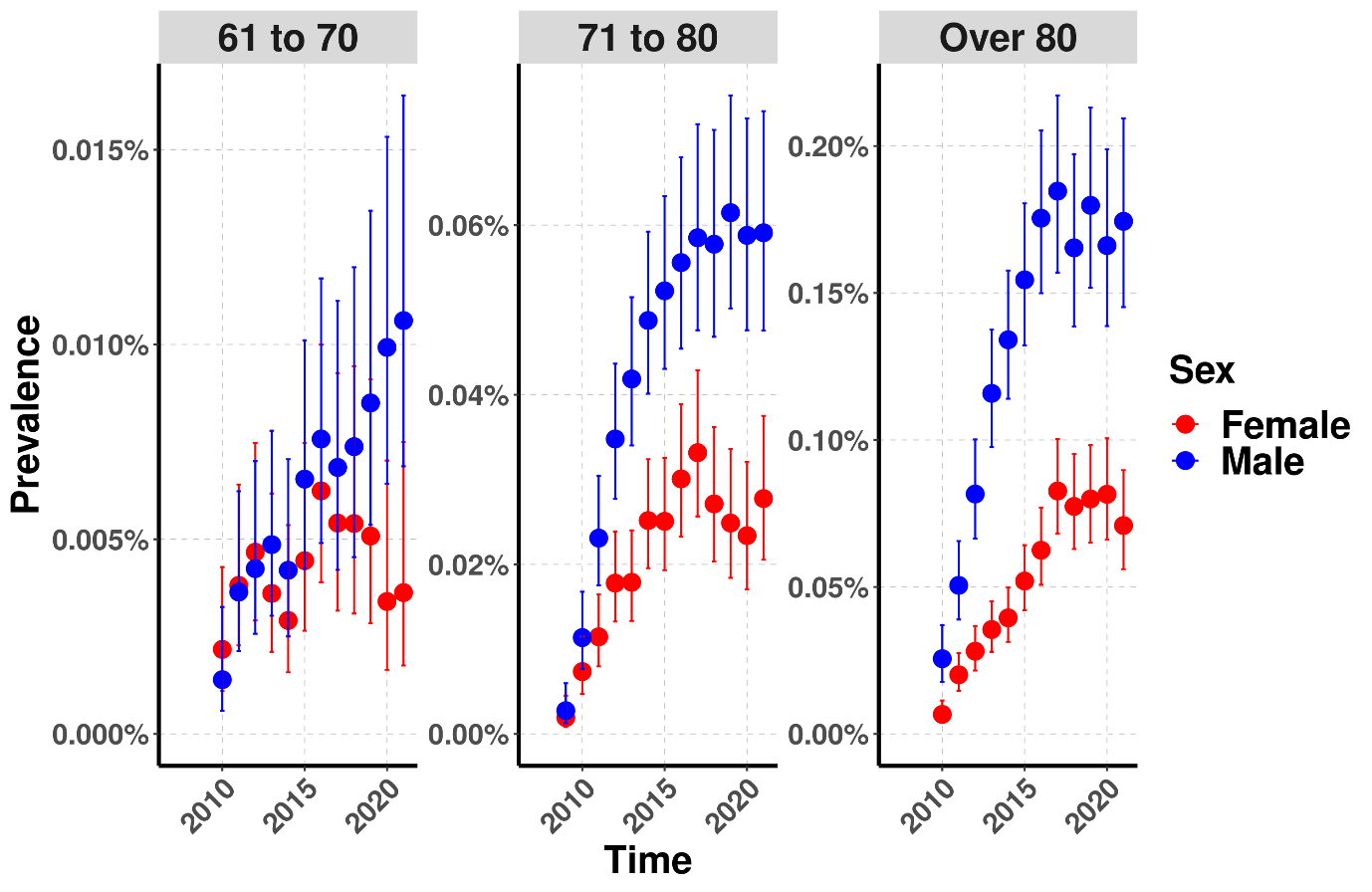

**Supplementary Figure 15: Annual prevalence of VP stratified by age and sex from 2007 to 2021**

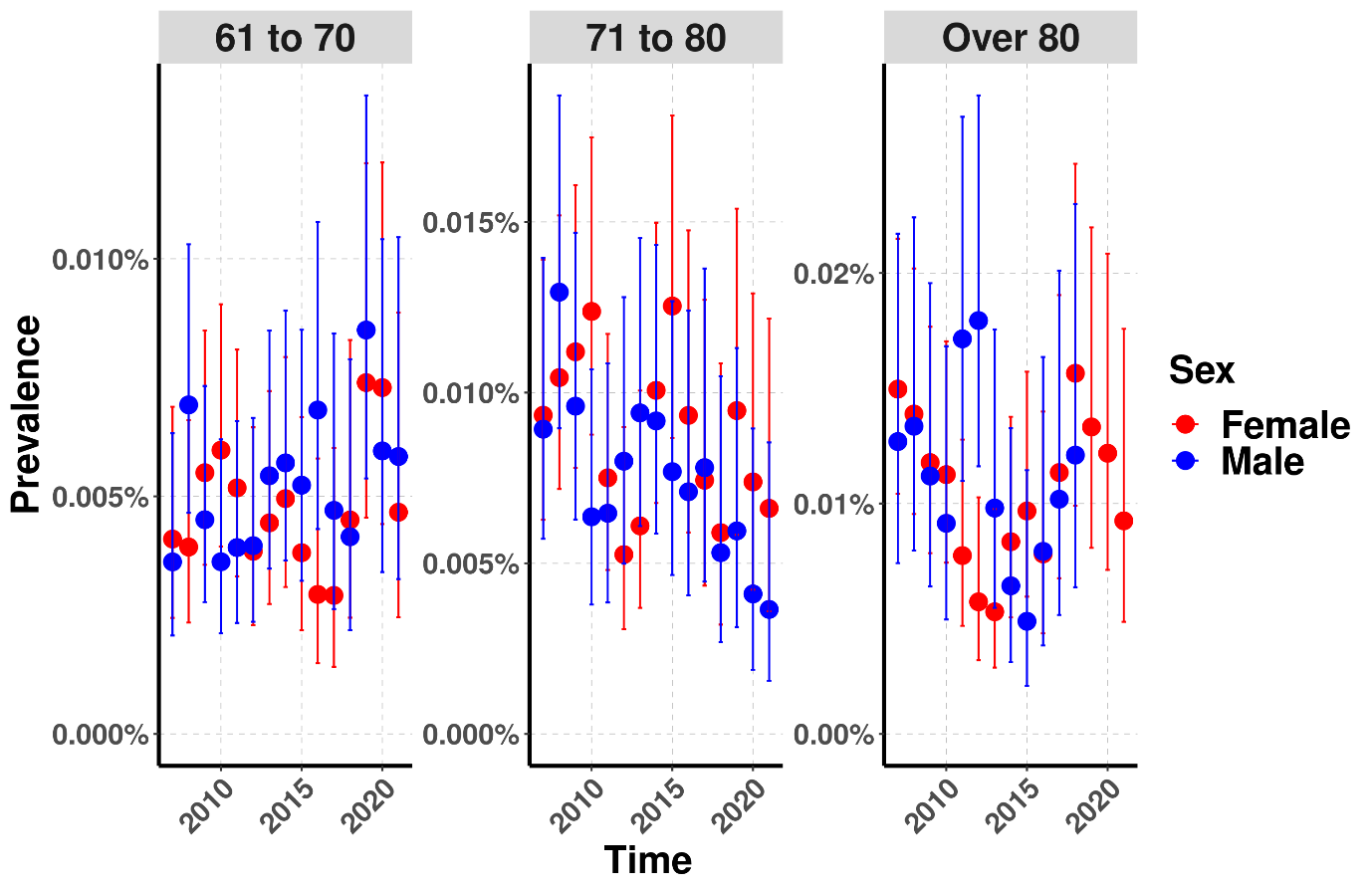

**Supplementary Figure 16: Annual prevalence of DIP stratified by age and sex from 2007 to 2021**

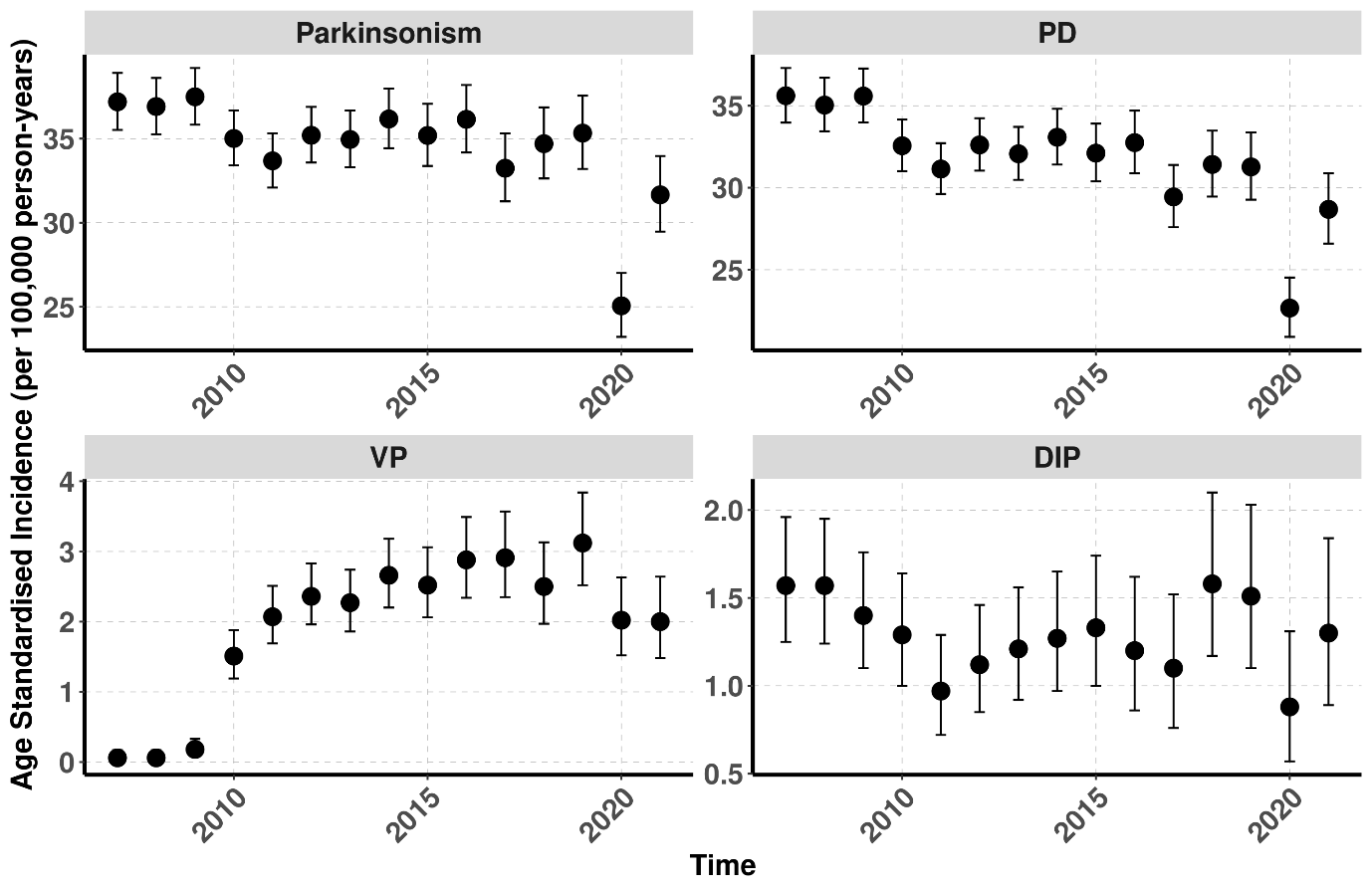

**Supplementary Figure 17: Age-standardized annual incidence of Parkinsonism and subtypes from 2007 to 2021**

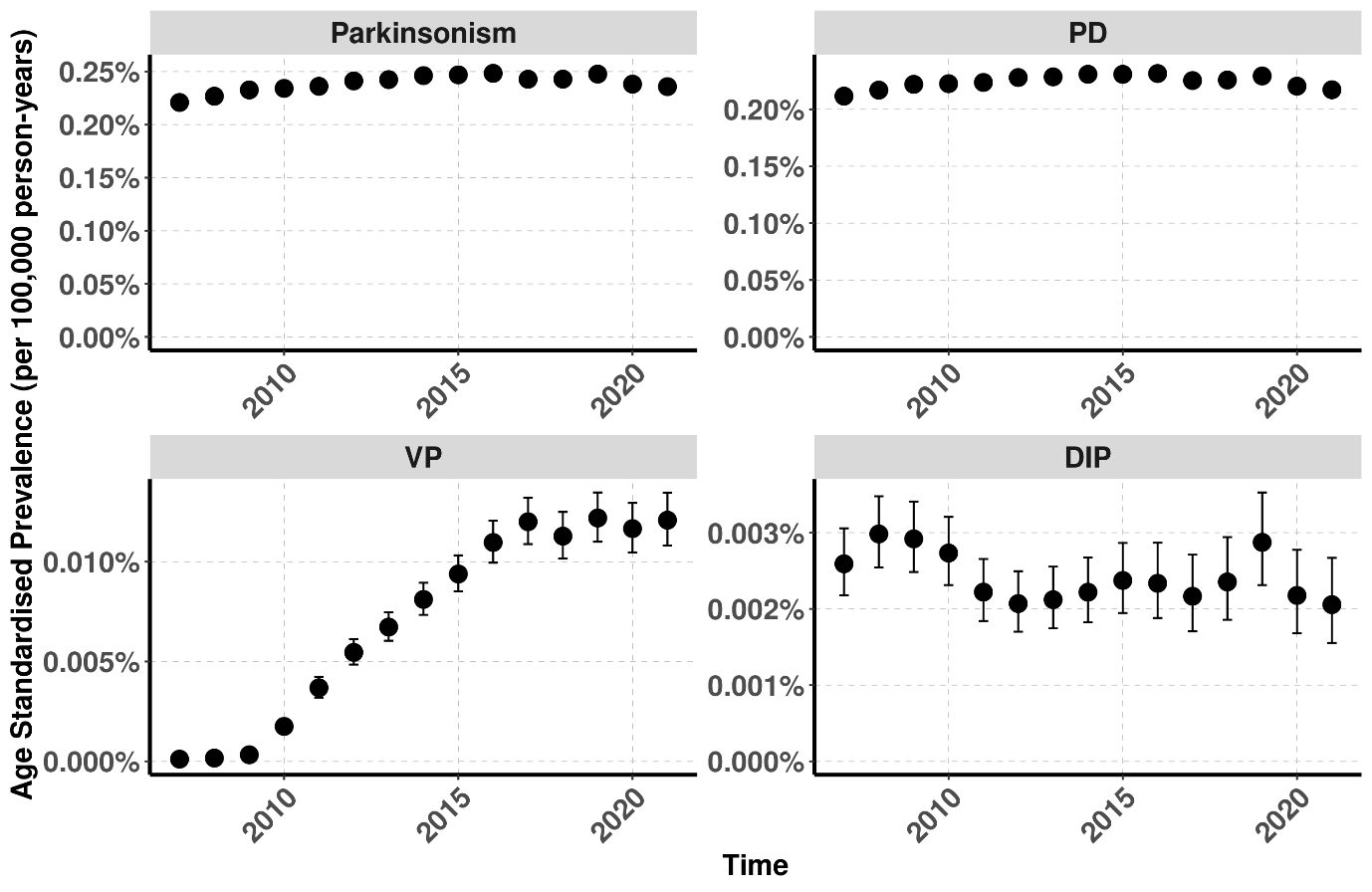
**Supplementary Figure 18: Age-standardized annual prevalence of Parkinsonism and subtypes from 2007 to 2021**

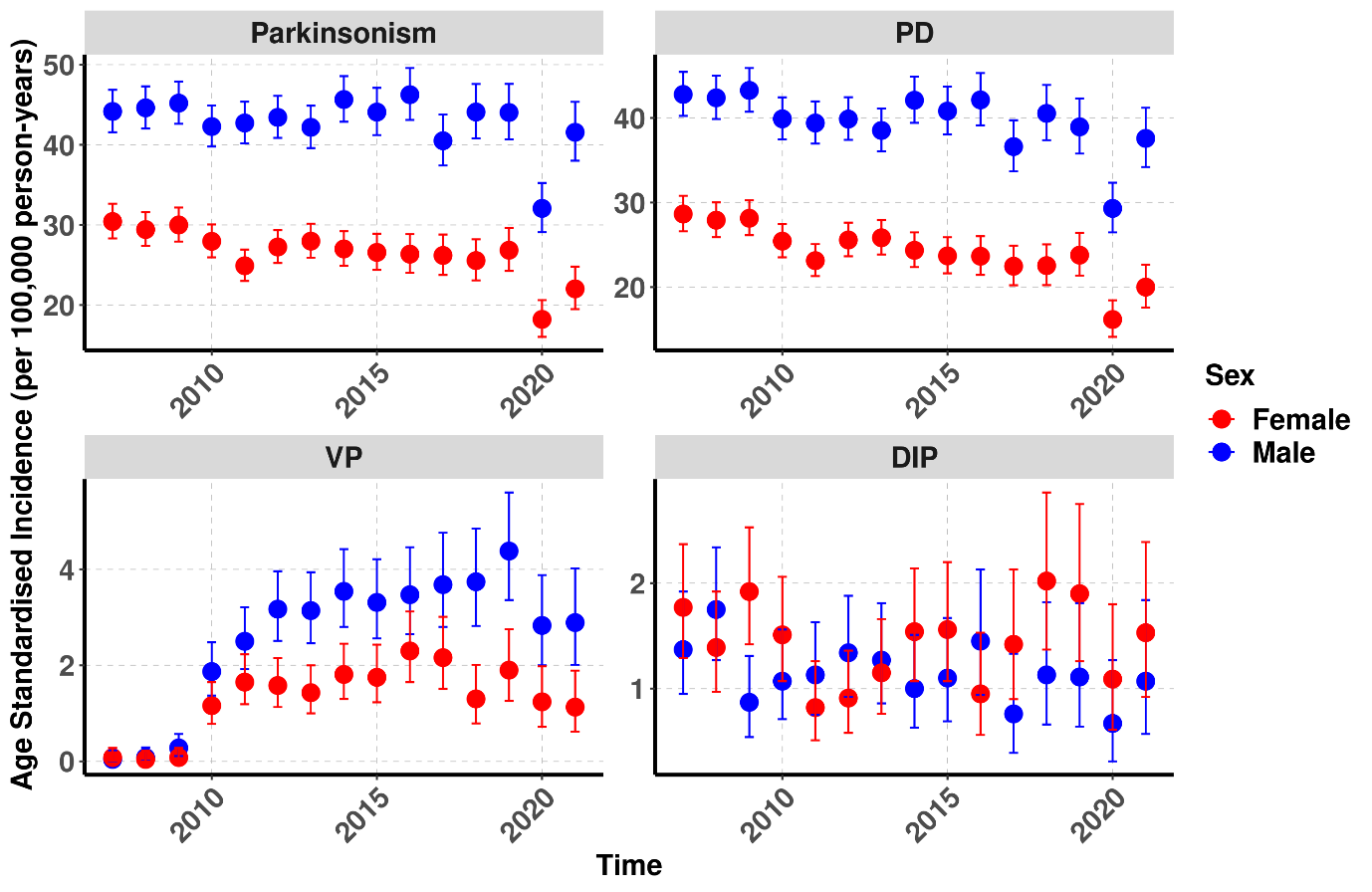

**Supplementary Figure 19: Age-standardized annual incidence of Parkinsonism and subtypes stratified by sex from 2007 to 2021**

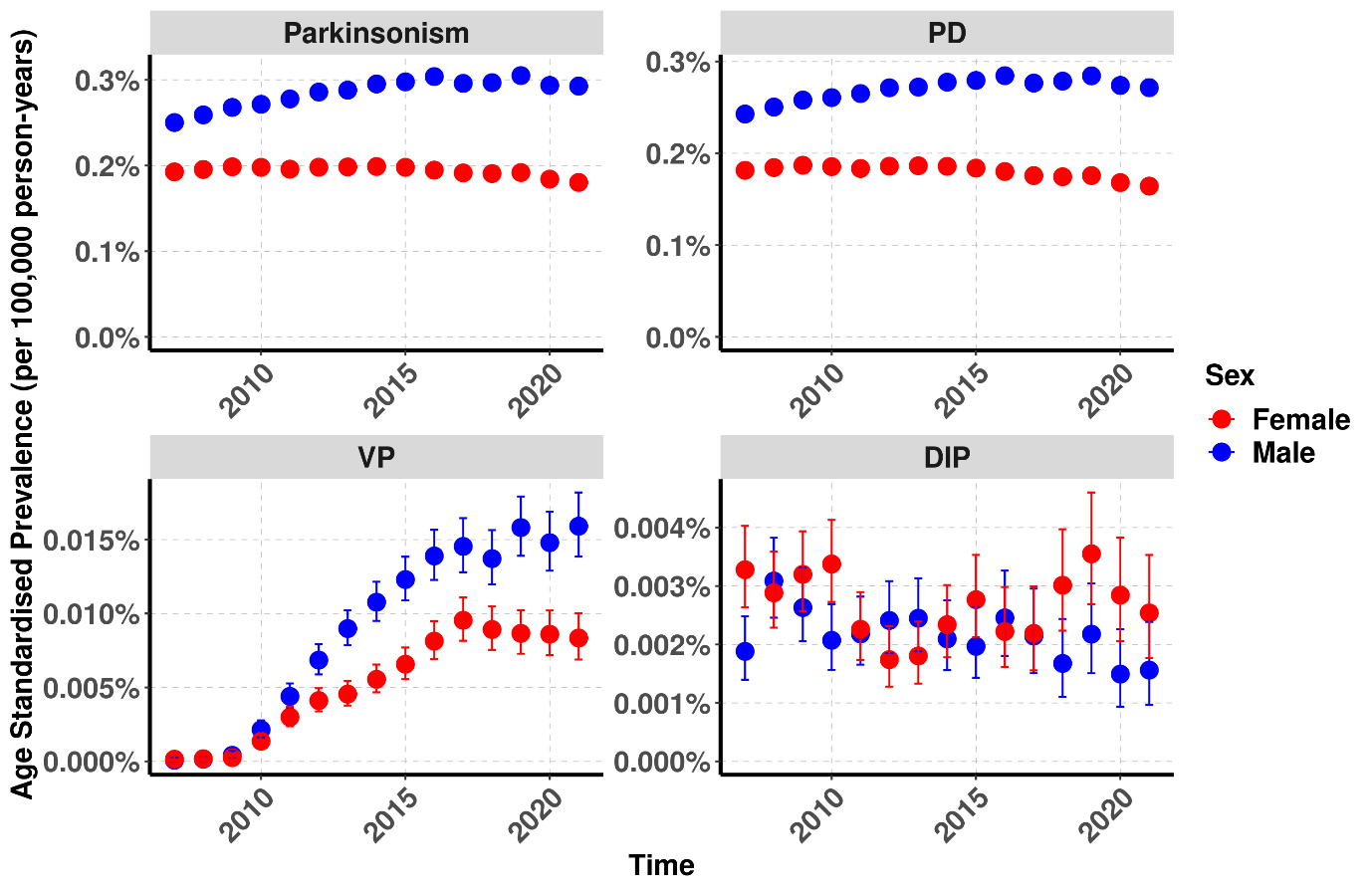
 **Supplementary Figure 20: Age-standardized annual prevalence of Parkinsonism and subtypes stratified by sex from 2007 to 2021**
