## supplementary_tables for "Secular trends of incidence and prevalence of parkinsonism and subtypes: A cohort study in the United Kingdom"

Catalogue:

| Supplement Table S1 | Condition codelists used to define outcomes of interest |
| --- | --- |
| Supplement Table S2 | Attrition table of each large-scale baseline characterization analysis |
| Supplement Table S3 | Age-standardized incidence of parkinsonism and subtypes and then stratified by sex |
| Supplement Table S4 | Age-standardized prevalence of parkinsonism and subtypes and then stratified by sex |

Supplementary Table S1: Condition codelists used to define outcomes of interest

The following codes have been used to carry out incidence and prevalence analysis for each outcome of interest. Only diagnosis codes are used for this study and these diagnosis codes are all SNOMED CT codes. These SNOMED CT codes are then mapped to OMOP CDM and the Concept ID column below is the correspoding concept ID in OMOP format. We developed concept definitions using ATLAS, the OHDSI open-source platform (https://github.com/OHDSI/atlas). Clinicians with primary care and neurology expertise further reviewed and refined these clinical codes.

| Concept ID | Name | Class | Domain | Vocabulary | Outcome of interest |
| --- | --- | --- | --- | --- | --- |
| 36715010 | Adult-onset dystonia parkinsonism | Disorder | Condition | SNOMED | Parkinsonism |
| 3654598 | Amyotrophic lateral sclerosis, parkinsonism, dementia complex | Disorder | Condition | SNOMED | Parkinsonism |
| 37166121 | Amyotrophic lateral sclerosis, parkinsonism, dementia complex of Kii Peninsula | Disorder | Condition | SNOMED | Parkinsonism |
| 37166119 | Amyotrophic lateral sclerosis, parkinsonism, dementia complex of West New Guinea | Disorder | Condition | SNOMED | Parkinsonism |
| 37166120 | Amyotrophic lateral sclerosis with parkinsonism | Disorder | Condition | SNOMED | Parkinsonism |
| 37312166 | ATPase cation transporting 13A2 related juvenile neuronal ceroid lipofuscinosis | Disorder | Condition | SNOMED | Parkinsonism |
| 37110776 | Atypical juvenile parkinsonism | Disorder | Condition | SNOMED | Parkinsonism |
| 36716783 | Atypical Parkinsonism | Disorder | Condition | SNOMED | Parkinsonism |
| 37396747 | Autosomal dominant late onset Parkinson disease | Disorder | Condition | SNOMED | Parkinsonism |
| 37110872 | Autosomal dominant striatal neurodegeneration | Disorder | Condition | SNOMED | Parkinsonism |
| 608078 | Autosomal recessive familial Parkinson disease | Disorder | Condition | SNOMED | Parkinsonism |
| 4046092 | Carbon monoxide-induced parkinsonism | Disorder | Condition | SNOMED | Parkinsonism |
| 4231949 | Cerebral degeneration due to Parkinson's disease | Disorder | Condition | SNOMED | Parkinsonism |
| 44792293 | Cerebral degeneration in Parkinson's disease | Disorder | Condition | SNOMED | Parkinsonism |
| 37164298 | Corticobasal syndrome | Disorder | Condition | SNOMED | Parkinsonism |
| 4314734 | Dementia associated with Parkinson's Disease | Disorder | Condition | SNOMED | Parkinsonism |
| 44782422 | Dementia due to Parkinson's disease | Disorder | Condition | SNOMED | Parkinsonism |
| 37311987 | Dissociative neurological symptom disorder co-occurrent with Parkinsonism | Disorder | Condition | SNOMED | Parkinsonism |
| 36716653 | Dopamine transporter deficiency syndrome | Disorder | Condition | SNOMED | Parkinsonism |
| 36716432 | Dystonia 16 | Disorder | Condition | SNOMED | Parkinsonism |
| 37399497 | Early onset parkinsonism and intellectual disability syndrome | Disorder | Condition | SNOMED | Parkinsonism |
| 1340429 | Exacerbation of Parkinsonism | Disorder | Condition | OMOP Extension | Parkinsonism |
| 1340428 | Exacerbation of Parkinson's disease | Disorder | Condition | OMOP Extension | Parkinsonism |
| 37163190 | Familial infantile bilateral striatal necrosis | Disorder | Condition | SNOMED | Parkinsonism |
| 37166883 | Familial multiple system tauopathy | Disorder | Condition | SNOMED | Parkinsonism |
| 45765480 | Frontotemporal dementia with parkinsonism-17 | Disorder | Condition | SNOMED | Parkinsonism |
| 37110549 | Functional parkinsonism | Disorder | Condition | SNOMED | Parkinsonism |
| 42537905 | Hemiparkinsonism hemiatrophy syndrome | Disorder | Condition | SNOMED | Parkinsonism |
| 37397519 | Infantile striatonigral degeneration | Disorder | Condition | SNOMED | Parkinsonism |
| 37117203 | Infection causing parkinsonism | Disorder | Condition | SNOMED | Parkinsonism |
| 4184466 | Juvenile paralysis agitans of Hunt | Disorder | Condition | SNOMED | Parkinsonism |
| 4047751 | Juvenile Parkinson's disease | Disorder | Condition | SNOMED | Parkinsonism |
| 37117179 | Kufor Rakeb syndrome | Disorder | Condition | SNOMED | Parkinsonism |
| 4044052 | Manganese-induced parkinsonism | Disorder | Condition | SNOMED | Parkinsonism |
| 4049300 | MPTP-induced parkinsonism | Disorder | Condition | SNOMED | Parkinsonism |
| 4248716 | Neuroleptic-induced parkinsonism | Disorder | Condition | SNOMED | Parkinsonism |
| 37164406 | Off-periods in Parkinson disease not responding to oral treatment | Disorder | Condition | SNOMED | Parkinsonism |
| 4142980 | On - off phenomenon | Disorder | Condition | SNOMED | Parkinsonism |
| 36713737 | Orthostatic hypotension co-occurrent and due to Parkinson's disease | Disorder | Condition | SNOMED | Parkinsonism |
| 37204371 | Parkinsonian pyramidal syndrome | Disorder | Condition | SNOMED | Parkinsonism |
| 4219273 | Parkinsonian syndrome with idiopathic orthostatic hypotension | Disorder | Condition | SNOMED | Parkinsonism |
| 4140090 | Parkinsonism | Disorder | Condition | SNOMED | Parkinsonism |
| 3655832 | Parkinsonism caused by carbon disulfide | Disorder | Condition | SNOMED | Parkinsonism |
| 35624223 | Parkinsonism caused by cyanide | Disorder | Condition | SNOMED | Parkinsonism |
| 37166926 | Parkinsonism caused by dopamine depleting agent | Disorder | Condition | SNOMED | Parkinsonism |
| 37166924 | Parkinsonism caused by dopamine receptor antagonist | Disorder | Condition | SNOMED | Parkinsonism |
| 3655826 | Parkinsonism caused by methanol | Disorder | Condition | SNOMED | Parkinsonism |
| 36716523 | Parkinsonism co-occurrent and due to acute infection | Disorder | Condition | SNOMED | Parkinsonism |
| 37110500 | Parkinsonism due to and following injury of head | Disorder | Condition | SNOMED | Parkinsonism |
| 4171569 | Parkinsonism due to drug | Disorder | Condition | SNOMED | Parkinsonism |
| 36716557 | Parkinsonism due to hereditary spastic paraplegia | Disorder | Condition | SNOMED | Parkinsonism |
| 36716784 | Parkinsonism due to heredodegenerative disorder | Disorder | Condition | SNOMED | Parkinsonism |
| 36716524 | Parkinsonism due to human immunodeficiency virus infection | Disorder | Condition | SNOMED | Parkinsonism |
| 37110501 | Parkinsonism due to mass lesion of brain | Disorder | Condition | SNOMED | Parkinsonism |
| 37161096 | Parkinsonism due to prion disease | Disorder | Condition | SNOMED | Parkinsonism |
| 36716525 | Parkinsonism following infection | Disorder | Condition | SNOMED | Parkinsonism |
| 37161098 | Parkinsonism following Mycoplasma infection | Disorder | Condition | SNOMED | Parkinsonism |
| 4043380 | Parkinsonism with calcification of basal ganglia | Disorder | Condition | SNOMED | Parkinsonism |
| 37396063 | Parkinsonism with dementia of Guadeloupe | Disorder | Condition | SNOMED | Parkinsonism |
| 381270 | Parkinson's disease | Disorder | Condition | SNOMED | Parkinsonism |
| 44783137 | Perry syndrome | Disorder | Condition | SNOMED | Parkinsonism |
| 37162109 | Pesticide-induced Parkinsonism | Disorder | Condition | SNOMED | Parkinsonism |
| 4064308 | Postencephalitic parkinsonism | Disorder | Condition | SNOMED | Parkinsonism |
| 608033 | Progressive supranuclear palsy parkinsonism syndrome | Disorder | Condition | SNOMED | Parkinsonism |
| 36714473 | Psychosis co-occurrent and due to Parkinson's disease | Disorder | Condition | SNOMED | Parkinsonism |
| 45765396 | Rapid onset dystonia parkinsonism | Disorder | Condition | SNOMED | Parkinsonism |
| 46269696 | Restrictive lung disease due to Parkinson disease | Disorder | Condition | SNOMED | Parkinsonism |
| 374013 | Secondary parkinsonism | Disorder | Condition | SNOMED | Parkinsonism |
| 605217 | Sporadic infantile bilateral striatal necrosis | Disorder | Condition | SNOMED | Parkinsonism |
| 37110499 | Sporadic Parkinson disease | Disorder | Condition | SNOMED | Parkinsonism |
| 4106410 | Striatonigral degeneration | Disorder | Condition | SNOMED | Parkinsonism |
| 4140881 | Symptomatic parkinsonism | Disorder | Condition | SNOMED | Parkinsonism |
| 373139 | Syphilitic parkinsonism | Disorder | Condition | SNOMED | Parkinsonism |
| 37162137 | Toxin-induced parkinsonism | Disorder | Condition | SNOMED | Parkinsonism |
| 4046093 | Vascular parkinsonism | Disorder | Condition | SNOMED | Parkinsonism |
| 44784241 | X-linked dystonia parkinsonism | Disorder | Condition | SNOMED | Parkinsonism |
| 36674193 | X-linked parkinsonism with spasticity syndrome | Disorder | Condition | SNOMED | Parkinsonism |
| 37395785 | Young onset Parkinson disease | Disorder | Condition | SNOMED | Parkinsonism |
| 4265907 | Parkinson-dementia complex of Guam | Disorder | Condition | SNOMED | Parkinsonism |
| 37110776 | Atypical juvenile parkinsonism | Disorder | Condition | SNOMED | PD |
| 37396747 | Autosomal dominant late onset Parkinson disease | Disorder | Condition | SNOMED | PD |
| 608078 | Autosomal recessive familial Parkinson disease | Disorder | Condition | SNOMED | PD |
| 4231949 | Cerebral degeneration due to Parkinson's disease | Disorder | Condition | SNOMED | PD |
| 44792293 | Cerebral degeneration in Parkinson's disease | Disorder | Condition | SNOMED | PD |
| 4314734 | Dementia associated with Parkinson's Disease | Disorder | Condition | SNOMED | PD |
| 44782422 | Dementia due to Parkinson's disease | Disorder | Condition | SNOMED | PD |
| 37399497 | Early onset parkinsonism and intellectual disability syndrome | Disorder | Condition | SNOMED | PD |
| 1340428 | Exacerbation of Parkinson's disease | Disorder | Condition | OMOP Extension | PD |
| 4184466 | Juvenile paralysis agitans of Hunt | Disorder | Condition | SNOMED | PD |
| 4047751 | Juvenile Parkinson's disease | Disorder | Condition | SNOMED | PD |
| 37164406 | Off-periods in Parkinson disease not responding to oral treatment | Disorder | Condition | SNOMED | PD |
| 36713737 | Orthostatic hypotension co-occurrent and due to Parkinson's disease | Disorder | Condition | SNOMED | PD |
| 37204371 | Parkinsonian pyramidal syndrome | Disorder | Condition | SNOMED | PD |
| 36716784 | Parkinsonism due to heredodegenerative disorder | Disorder | Condition | SNOMED | PD |
| 4043380 | Parkinsonism with calcification of basal ganglia | Disorder | Condition | SNOMED | PD |
| 381270 | Parkinson's disease | Disorder | Condition | SNOMED | PD |
| 36714473 | Psychosis co-occurrent and due to Parkinson's disease | Disorder | Condition | SNOMED | PD |
| 46269696 | Restrictive lung disease due to Parkinson disease | Disorder | Condition | SNOMED | PD |
| 37110499 | Sporadic Parkinson disease | Disorder | Condition | SNOMED | PD |
| 37395785 | Young onset Parkinson disease | Disorder | Condition | SNOMED | PD |
| 4265907 | Parkinson-dementia complex of Guam | Disorder | Condition | SNOMED | PD |
| 4046093 | Vascular parkinsonism | Disorder | Condition | SNOMED | VP |
| 4248716 | Neuroleptic-induced parkinsonism | Disorder | Condition | SNOMED | DIP |
| 4142980 | On - off phenomenon | Disorder | Condition | SNOMED | DIP |
| 37166926 | Parkinsonism caused by dopamine depleting agent | Disorder | Condition | SNOMED | DIP |
| 37166924 | Parkinsonism caused by dopamine receptor antagonist | Disorder | Condition | SNOMED | DIP |
| 3655826 | Parkinsonism caused by methanol | Disorder | Condition | SNOMED | DIP |
| 4171569 | Parkinsonism due to drug | Disorder | Condition | SNOMED | DIP |

Supplementary Table S2: Attrition table of each large-scale baseline characterization analysis

| **N** | **Reason** | **N excluded** | **Parkinsonism and subtypes** |
| --- | --- | --- | --- |
| 17054819 | Starting population | NA | Parkinsonism |
| 17054819 | Missing year of birth | 0 |  |
| 17054819 | Missing sex | 0 |  |
| 15210165 | Cannot satisfy age criteria during the study period based on year of birth | 1844654 |  |
| 11246146 | No observation time available during study period | 3964019 |  |
| 11246146 | Doesn't satisfy age criteria during the study period | 0 |  |
| 10150486 | Prior history requirement not fulfilled during study period | 1095660 |  |
| 9604592 | No observation time available after applying age and prior history criteria | 545894 |  |
| 9639215 | Starting analysis population | NA |  |
| 9591649 | Excluded due to prior event (do not pass outcome washout during study period) | 47566 |  |
| 9591649 | Not observed during the complete database interval | 0 |  |
| 17054819 | Starting population | NA | Drug-induced Parkinsonism |
| 17054819 | Missing year of birth | 0 |  |
| 17054819 | Missing sex | 0 |  |
| 15210165 | Cannot satisfy age criteria during the study period based on year of birth | 1844654 |  |
| 11246146 | No observation time available during study period | 3964019 |  |
| 11246146 | Doesn't satisfy age criteria during the study period | 0 |  |
| 10150486 | Prior history requirement not fulfilled during study period | 1095660 |  |
| 9604592 | No observation time available after applying age and prior history criteria | 545894 |  |
| 9605882 | Starting analysis population | NA |  |
| 9604111 | Excluded due to prior event (do not pass outcome washout during study period) | 1771 |  |
| 9604111 | Not observed during the complete database interval | 0 |  |
| 17054819 | Starting population | NA | Parkinson’s Disease |
| 17054819 | Missing year of birth | 0 |  |
| 17054819 | Missing sex | 0 |  |
| 15210165 | Cannot satisfy age criteria during the study period based on year of birth | 1844654 |  |
| 11246146 | No observation time available during study period | 3964019 |  |
| 11246146 | Doesn't satisfy age criteria during the study period | 0 |  |
| 10150486 | Prior history requirement not fulfilled during study period | 1095660 |  |
| 9604592 | No observation time available after applying age and prior history criteria | 545894 |  |
| 9636979 | Starting analysis population | NA |  |
| 9592211 | Excluded due to prior event (do not pass outcome washout during study period) | 44768 |  |
| 9592211 | Not observed during the complete database interval | 0 |  |
| 17054819 | Starting population | NA | Vascular Parkinsonism |
| 17054819 | Missing year of birth | 0 |  |
| 17054819 | Missing sex | 0 |  |
| 15210165 | Cannot satisfy age criteria during the study period based on year of birth | 1844654 |  |
| 11246146 | No observation time available during study period | 3964019 |  |
| 11246146 | Doesn't satisfy age criteria during the study period | 0 |  |
| 10150486 | Prior history requirement not fulfilled during study period | 1095660 |  |
| 9604592 | No observation time available after applying age and prior history criteria | 545894 |  |
| 9605804 | Starting analysis population | NA |  |
| 9604509 | Excluded due to prior event (do not pass outcome washout during study period) | 1295 |  |
| 9604509 | Not observed during the complete database interval | 0 |  |

Supplementary Table S3: Age-standardized incidence of parkinsonism and subtypes and then stratified by sex

| incidence_start_date | incidence_end_date | n_events | person-years | standardised_incidence | standardised_incidence_lower | standardised_incidence_upper | outcome_cohort_name | denominator_sex |
| --- | --- | --- | --- | --- | --- | --- | --- | --- |
| 01/01/2007 | 31/12/2007 | 79 | 5019494.267 | 1.57 | 1.25 | 1.96 | DrugInducedParkinsonism | Both |
| 01/01/2008 | 31/12/2008 | 80 | 5111792.309 | 1.57 | 1.24 | 1.95 | DrugInducedParkinsonism | Both |
| 01/01/2009 | 31/12/2009 | 72 | 5139828.287 | 1.4 | 1.1 | 1.76 | DrugInducedParkinsonism | Both |
| 01/01/2010 | 31/12/2010 | 66 | 5105412.099 | 1.29 | 1 | 1.64 | DrugInducedParkinsonism | Both |
| 01/01/2011 | 31/12/2011 | 49 | 5025774.53 | 0.97 | 0.72 | 1.29 | DrugInducedParkinsonism | Both |
| 01/01/2012 | 31/12/2012 | 56 | 4992802.166 | 1.12 | 0.85 | 1.46 | DrugInducedParkinsonism | Both |
| 01/01/2013 | 31/12/2013 | 58 | 4799260.747 | 1.21 | 0.92 | 1.56 | DrugInducedParkinsonism | Both |
| 01/01/2014 | 31/12/2014 | 57 | 4471378.842 | 1.27 | 0.97 | 1.65 | DrugInducedParkinsonism | Both |
| 01/01/2015 | 31/12/2015 | 54 | 4046795.034 | 1.33 | 1 | 1.74 | DrugInducedParkinsonism | Both |
| 01/01/2016 | 31/12/2016 | 42 | 3512697.175 | 1.2 | 0.86 | 1.62 | DrugInducedParkinsonism | Both |
| 01/01/2017 | 31/12/2017 | 35 | 3192565.306 | 1.1 | 0.76 | 1.52 | DrugInducedParkinsonism | Both |
| 01/01/2018 | 31/12/2018 | 48 | 3035761.67 | 1.58 | 1.17 | 2.1 | DrugInducedParkinsonism | Both |
| 01/01/2019 | 31/12/2019 | 44 | 2913047.318 | 1.51 | 1.1 | 2.03 | DrugInducedParkinsonism | Both |
| 01/01/2020 | 31/12/2020 | 24 | 2719635.871 | 0.88 | 0.57 | 1.31 | DrugInducedParkinsonism | Both |
| 01/01/2021 | 31/12/2021 | 32 | 2452629.643 | 1.3 | 0.89 | 1.84 | DrugInducedParkinsonism | Both |
| 01/01/2007 | 31/12/2007 | 1863 | 5009900.367 | 37.19 | 35.52 | 38.91 | Parkinsonism | Both |
| 01/01/2008 | 31/12/2008 | 1883 | 5101785.443 | 36.91 | 35.26 | 38.61 | Parkinsonism | Both |
| 01/01/2009 | 31/12/2009 | 1923 | 5129539.967 | 37.49 | 35.83 | 39.2 | Parkinsonism | Both |
| 01/01/2010 | 31/12/2010 | 1784 | 5095019.526 | 35.01 | 33.41 | 36.68 | Parkinsonism | Both |
| 01/01/2011 | 31/12/2011 | 1689 | 5015340.709 | 33.68 | 32.09 | 35.32 | Parkinsonism | Both |
| 01/01/2012 | 31/12/2012 | 1754 | 4982335.354 | 35.2 | 33.58 | 36.89 | Parkinsonism | Both |
| 01/01/2013 | 31/12/2013 | 1674 | 4789109.15 | 34.95 | 33.3 | 36.67 | Parkinsonism | Both |
| 01/01/2014 | 31/12/2014 | 1614 | 4461744.986 | 36.17 | 34.43 | 37.98 | Parkinsonism | Both |
| 01/01/2015 | 31/12/2015 | 1421 | 4038073.949 | 35.19 | 33.38 | 37.07 | Parkinsonism | Both |
| 01/01/2016 | 31/12/2016 | 1267 | 3505135.039 | 36.15 | 34.18 | 38.19 | Parkinsonism | Both |
| 01/01/2017 | 31/12/2017 | 1059 | 3185745.886 | 33.24 | 31.27 | 35.31 | Parkinsonism | Both |
| 01/01/2018 | 31/12/2018 | 1051 | 3029234.305 | 34.7 | 32.63 | 36.86 | Parkinsonism | Both |
| 01/01/2019 | 31/12/2019 | 1027 | 2906701.572 | 35.33 | 33.2 | 37.56 | Parkinsonism | Both |
| 01/01/2020 | 31/12/2020 | 680 | 2713905.908 | 25.06 | 23.21 | 27.01 | Parkinsonism | Both |
| 01/01/2021 | 31/12/2021 | 775 | 2447522.483 | 31.66 | 29.47 | 33.97 | Parkinsonism | Both |
| 01/01/2007 | 31/12/2007 | 1784 | 5010327.767 | 35.61 | 33.97 | 37.3 | ParkinsonsDisease | Both |
| 01/01/2008 | 31/12/2008 | 1788 | 5102241.388 | 35.04 | 33.44 | 36.71 | ParkinsonsDisease | Both |
| 01/01/2009 | 31/12/2009 | 1826 | 5130033.826 | 35.59 | 33.98 | 37.27 | ParkinsonsDisease | Both |
| 01/01/2010 | 31/12/2010 | 1659 | 5095537.692 | 32.56 | 31.01 | 34.16 | ParkinsonsDisease | Both |
| 01/01/2011 | 31/12/2011 | 1562 | 5015895.039 | 31.14 | 29.62 | 32.72 | ParkinsonsDisease | Both |
| 01/01/2012 | 31/12/2012 | 1625 | 4982923.951 | 32.61 | 31.04 | 34.24 | ParkinsonsDisease | Both |
| 01/01/2013 | 31/12/2013 | 1536 | 4789704.06 | 32.07 | 30.48 | 33.71 | ParkinsonsDisease | Both |
| 01/01/2014 | 31/12/2014 | 1476 | 4462355.723 | 33.08 | 31.41 | 34.81 | ParkinsonsDisease | Both |
| 01/01/2015 | 31/12/2015 | 1297 | 4038651.157 | 32.11 | 30.39 | 33.91 | ParkinsonsDisease | Both |
| 01/01/2016 | 31/12/2016 | 1148 | 3505646.355 | 32.75 | 30.88 | 34.7 | ParkinsonsDisease | Both |
| 01/01/2017 | 31/12/2017 | 938 | 3186220.025 | 29.44 | 27.59 | 31.39 | ParkinsonsDisease | Both |
| 01/01/2018 | 31/12/2018 | 952 | 3029680.756 | 31.42 | 29.46 | 33.48 | ParkinsonsDisease | Both |
| 01/01/2019 | 31/12/2019 | 909 | 2907162.275 | 31.27 | 29.27 | 33.37 | ParkinsonsDisease | Both |
| 01/01/2020 | 31/12/2020 | 615 | 2714347.551 | 22.66 | 20.9 | 24.52 | ParkinsonsDisease | Both |
| 01/01/2021 | 31/12/2021 | 702 | 2447924.742 | 28.68 | 26.59 | 30.88 | ParkinsonsDisease | Both |
| 01/01/2007 | 31/12/2007 | 3 | 5019905.884 | 0.06 | 0.01 | 0.17 | VascularParkinsonism | Both |
| 01/01/2008 | 31/12/2008 | 3 | 5112220.893 | 0.06 | 0.01 | 0.17 | VascularParkinsonism | Both |
| 01/01/2009 | 31/12/2009 | 9 | 5140268.767 | 0.18 | 0.08 | 0.33 | VascularParkinsonism | Both |
| 01/01/2010 | 31/12/2010 | 77 | 5105819.762 | 1.51 | 1.19 | 1.88 | VascularParkinsonism | Both |
| 01/01/2011 | 31/12/2011 | 104 | 5026074.062 | 2.07 | 1.69 | 2.51 | VascularParkinsonism | Both |
| 01/01/2012 | 31/12/2012 | 118 | 4993033.265 | 2.36 | 1.96 | 2.83 | VascularParkinsonism | Both |
| 01/01/2013 | 31/12/2013 | 109 | 4799416.11 | 2.27 | 1.86 | 2.74 | VascularParkinsonism | Both |
| 01/01/2014 | 31/12/2014 | 119 | 4471463.992 | 2.66 | 2.2 | 3.18 | VascularParkinsonism | Both |
| 01/01/2015 | 31/12/2015 | 102 | 4046823.814 | 2.52 | 2.06 | 3.06 | VascularParkinsonism | Both |
| 01/01/2016 | 31/12/2016 | 101 | 3512640.402 | 2.88 | 2.34 | 3.49 | VascularParkinsonism | Both |
| 01/01/2017 | 31/12/2017 | 93 | 3192492.665 | 2.91 | 2.35 | 3.57 | VascularParkinsonism | Both |
| 01/01/2018 | 31/12/2018 | 76 | 3035707.296 | 2.5 | 1.97 | 3.13 | VascularParkinsonism | Both |
| 01/01/2019 | 31/12/2019 | 91 | 2912975.559 | 3.12 | 2.52 | 3.84 | VascularParkinsonism | Both |
| 01/01/2020 | 31/12/2020 | 55 | 2719573.35 | 2.02 | 1.52 | 2.63 | VascularParkinsonism | Both |
| 01/01/2021 | 31/12/2021 | 49 | 2452577.84 | 2 | 1.48 | 2.64 | VascularParkinsonism | Both |
| 01/01/2007 | 31/12/2007 | 34 | 2473839.488 | 1.37 | 0.95 | 1.92 | DrugInducedParkinsonism | Male |
| 01/01/2008 | 31/12/2008 | 44 | 2520969.385 | 1.75 | 1.27 | 2.34 | DrugInducedParkinsonism | Male |
| 01/01/2009 | 31/12/2009 | 22 | 2534544.402 | 0.87 | 0.54 | 1.31 | DrugInducedParkinsonism | Male |
| 01/01/2010 | 31/12/2010 | 27 | 2517561.544 | 1.07 | 0.71 | 1.56 | DrugInducedParkinsonism | Male |
| 01/01/2011 | 31/12/2011 | 28 | 2475632.12 | 1.13 | 0.75 | 1.63 | DrugInducedParkinsonism | Male |
| 01/01/2012 | 31/12/2012 | 33 | 2458642.743 | 1.34 | 0.92 | 1.88 | DrugInducedParkinsonism | Male |
| 01/01/2013 | 31/12/2013 | 30 | 2360335.354 | 1.27 | 0.86 | 1.81 | DrugInducedParkinsonism | Male |
| 01/01/2014 | 31/12/2014 | 22 | 2200648.914 | 1 | 0.63 | 1.51 | DrugInducedParkinsonism | Male |
| 01/01/2015 | 31/12/2015 | 22 | 1992314.07 | 1.1 | 0.69 | 1.67 | DrugInducedParkinsonism | Male |
| 01/01/2016 | 31/12/2016 | 25 | 1729801.161 | 1.45 | 0.94 | 2.13 | DrugInducedParkinsonism | Male |
| 01/01/2017 | 31/12/2017 | 12 | 1574477.216 | 0.76 | 0.39 | 1.33 | DrugInducedParkinsonism | Male |
| 01/01/2018 | 31/12/2018 | 17 | 1498694.724 | 1.13 | 0.66 | 1.82 | DrugInducedParkinsonism | Male |
| 01/01/2019 | 31/12/2019 | 16 | 1439173.429 | 1.11 | 0.64 | 1.81 | DrugInducedParkinsonism | Male |
| 01/01/2020 | 31/12/2020 | 9 | 1344650.286 | 0.67 | 0.31 | 1.27 | DrugInducedParkinsonism | Male |
| 01/01/2021 | 31/12/2021 | 13 | 1210946.861 | 1.07 | 0.57 | 1.84 | DrugInducedParkinsonism | Male |
| 01/01/2007 | 31/12/2007 | 1090 | 2468462.371 | 44.16 | 41.57 | 46.86 | Parkinsonism | Male |
| 01/01/2008 | 31/12/2008 | 1122 | 2515295.069 | 44.61 | 42.03 | 47.3 | Parkinsonism | Male |
| 01/01/2009 | 31/12/2009 | 1143 | 2528662.5 | 45.2 | 42.62 | 47.9 | Parkinsonism | Male |
| 01/01/2010 | 31/12/2010 | 1062 | 2511554.91 | 42.28 | 39.78 | 44.91 | Parkinsonism | Male |
| 01/01/2011 | 31/12/2011 | 1055 | 2469551.143 | 42.72 | 40.18 | 45.38 | Parkinsonism | Male |
| 01/01/2012 | 31/12/2012 | 1065 | 2452509.774 | 43.42 | 40.86 | 46.11 | Parkinsonism | Male |
| 01/01/2013 | 31/12/2013 | 993 | 2354356.052 | 42.18 | 39.59 | 44.88 | Parkinsonism | Male |
| 01/01/2014 | 31/12/2014 | 1002 | 2194951.428 | 45.65 | 42.87 | 48.57 | Parkinsonism | Male |
| 01/01/2015 | 31/12/2015 | 876 | 1987101.867 | 44.08 | 41.21 | 47.1 | Parkinsonism | Male |
| 01/01/2016 | 31/12/2016 | 798 | 1725226.448 | 46.25 | 43.1 | 49.58 | Parkinsonism | Male |
| 01/01/2017 | 31/12/2017 | 636 | 1570360.246 | 40.5 | 37.41 | 43.77 | Parkinsonism | Male |
| 01/01/2018 | 31/12/2018 | 659 | 1494727.236 | 44.09 | 40.79 | 47.59 | Parkinsonism | Male |
| 01/01/2019 | 31/12/2019 | 632 | 1435281.487 | 44.03 | 40.67 | 47.6 | Parkinsonism | Male |
| 01/01/2020 | 31/12/2020 | 430 | 1341126.103 | 32.06 | 29.1 | 35.24 | Parkinsonism | Male |
| 01/01/2021 | 31/12/2021 | 502 | 1207805.64 | 41.56 | 38.01 | 45.36 | Parkinsonism | Male |
| 01/01/2007 | 31/12/2007 | 1056 | 2468624.194 | 42.78 | 40.24 | 45.44 | ParkinsonsDisease | Male |
| 01/01/2008 | 31/12/2008 | 1066 | 2515484.986 | 42.38 | 39.87 | 45 | ParkinsonsDisease | Male |
| 01/01/2009 | 31/12/2009 | 1094 | 2528882.491 | 43.26 | 40.73 | 45.9 | ParkinsonsDisease | Male |
| 01/01/2010 | 31/12/2010 | 1002 | 2511791.885 | 39.89 | 37.46 | 42.44 | ParkinsonsDisease | Male |
| 01/01/2011 | 31/12/2011 | 973 | 2469819.907 | 39.4 | 36.96 | 41.95 | ParkinsonsDisease | Male |
| 01/01/2012 | 31/12/2012 | 978 | 2452816.312 | 39.87 | 37.41 | 42.45 | ParkinsonsDisease | Male |
| 01/01/2013 | 31/12/2013 | 907 | 2354689.451 | 38.52 | 36.05 | 41.11 | ParkinsonsDisease | Male |
| 01/01/2014 | 31/12/2014 | 924 | 2195296.575 | 42.09 | 39.42 | 44.89 | ParkinsonsDisease | Male |
| 01/01/2015 | 31/12/2015 | 811 | 1987421.517 | 40.81 | 38.05 | 43.71 | ParkinsonsDisease | Male |
| 01/01/2016 | 31/12/2016 | 727 | 1725513.333 | 42.13 | 39.12 | 45.31 | ParkinsonsDisease | Male |
| 01/01/2017 | 31/12/2017 | 575 | 1570613.604 | 36.61 | 33.68 | 39.73 | ParkinsonsDisease | Male |
| 01/01/2018 | 31/12/2018 | 606 | 1494958.612 | 40.54 | 37.37 | 43.9 | ParkinsonsDisease | Male |
| 01/01/2019 | 31/12/2019 | 559 | 1435530.543 | 38.94 | 35.78 | 42.31 | ParkinsonsDisease | Male |
| 01/01/2020 | 31/12/2020 | 393 | 1341361.689 | 29.3 | 26.47 | 32.34 | ParkinsonsDisease | Male |
| 01/01/2021 | 31/12/2021 | 454 | 1208025.336 | 37.58 | 34.2 | 41.2 | ParkinsonsDisease | Male |
| 01/01/2007 | 31/12/2007 | 1 | 2473987.354 | 0.04 | 0 | 0.23 | VascularParkinsonism | Male |
| 01/01/2008 | 31/12/2008 | 2 | 2521137.971 | 0.08 | 0.01 | 0.29 | VascularParkinsonism | Male |
| 01/01/2009 | 31/12/2009 | 7 | 2534722.552 | 0.28 | 0.11 | 0.57 | VascularParkinsonism | Male |
| 01/01/2010 | 31/12/2010 | 47 | 2517714.349 | 1.87 | 1.37 | 2.48 | VascularParkinsonism | Male |
| 01/01/2011 | 31/12/2011 | 62 | 2475734.363 | 2.5 | 1.92 | 3.21 | VascularParkinsonism | Male |
| 01/01/2012 | 31/12/2012 | 78 | 2458707.146 | 3.17 | 2.51 | 3.96 | VascularParkinsonism | Male |
| 01/01/2013 | 31/12/2013 | 74 | 2360357.533 | 3.14 | 2.46 | 3.94 | VascularParkinsonism | Male |
| 01/01/2014 | 31/12/2014 | 78 | 2200626.453 | 3.54 | 2.8 | 4.42 | VascularParkinsonism | Male |
| 01/01/2015 | 31/12/2015 | 66 | 1992271.981 | 3.31 | 2.56 | 4.21 | VascularParkinsonism | Male |
| 01/01/2016 | 31/12/2016 | 60 | 1729722.686 | 3.47 | 2.65 | 4.46 | VascularParkinsonism | Male |
| 01/01/2017 | 31/12/2017 | 58 | 1574406.215 | 3.68 | 2.8 | 4.76 | VascularParkinsonism | Male |
| 01/01/2018 | 31/12/2018 | 56 | 1498631.762 | 3.74 | 2.82 | 4.85 | VascularParkinsonism | Male |
| 01/01/2019 | 31/12/2019 | 63 | 1439087.598 | 4.38 | 3.36 | 5.6 | VascularParkinsonism | Male |
| 01/01/2020 | 31/12/2020 | 38 | 1344565.32 | 2.83 | 2 | 3.88 | VascularParkinsonism | Male |
| 01/01/2021 | 31/12/2021 | 35 | 1210865.648 | 2.89 | 2.01 | 4.02 | VascularParkinsonism | Male |
| 01/01/2007 | 31/12/2007 | 45 | 2545654.779 | 1.77 | 1.29 | 2.37 | DrugInducedParkinsonism | Female |
| 01/01/2008 | 31/12/2008 | 36 | 2590822.924 | 1.39 | 0.97 | 1.92 | DrugInducedParkinsonism | Female |
| 01/01/2009 | 31/12/2009 | 50 | 2605283.885 | 1.92 | 1.42 | 2.53 | DrugInducedParkinsonism | Female |
| 01/01/2010 | 31/12/2010 | 39 | 2587850.554 | 1.51 | 1.07 | 2.06 | DrugInducedParkinsonism | Female |
| 01/01/2011 | 31/12/2011 | 21 | 2550142.409 | 0.82 | 0.51 | 1.26 | DrugInducedParkinsonism | Female |
| 01/01/2012 | 31/12/2012 | 23 | 2534159.422 | 0.91 | 0.58 | 1.36 | DrugInducedParkinsonism | Female |
| 01/01/2013 | 31/12/2013 | 28 | 2438925.394 | 1.15 | 0.76 | 1.66 | DrugInducedParkinsonism | Female |
| 01/01/2014 | 31/12/2014 | 35 | 2270729.927 | 1.54 | 1.07 | 2.14 | DrugInducedParkinsonism | Female |
| 01/01/2015 | 31/12/2015 | 32 | 2054480.964 | 1.56 | 1.07 | 2.2 | DrugInducedParkinsonism | Female |
| 01/01/2016 | 31/12/2016 | 17 | 1782896.014 | 0.95 | 0.56 | 1.53 | DrugInducedParkinsonism | Female |
| 01/01/2017 | 31/12/2017 | 23 | 1618088.09 | 1.42 | 0.9 | 2.13 | DrugInducedParkinsonism | Female |
| 01/01/2018 | 31/12/2018 | 31 | 1537066.946 | 2.02 | 1.37 | 2.86 | DrugInducedParkinsonism | Female |
| 01/01/2019 | 31/12/2019 | 28 | 1473873.889 | 1.9 | 1.26 | 2.75 | DrugInducedParkinsonism | Female |
| 01/01/2020 | 31/12/2020 | 15 | 1374985.585 | 1.09 | 0.61 | 1.8 | DrugInducedParkinsonism | Female |
| 01/01/2021 | 31/12/2021 | 19 | 1241682.782 | 1.53 | 0.92 | 2.39 | DrugInducedParkinsonism | Female |
| 01/01/2007 | 31/12/2007 | 773 | 2541437.996 | 30.42 | 28.31 | 32.64 | Parkinsonism | Female |
| 01/01/2008 | 31/12/2008 | 761 | 2586490.374 | 29.42 | 27.37 | 31.59 | Parkinsonism | Female |
| 01/01/2009 | 31/12/2009 | 780 | 2600877.467 | 29.99 | 27.92 | 32.17 | Parkinsonism | Female |
| 01/01/2010 | 31/12/2010 | 722 | 2583464.616 | 27.95 | 25.95 | 30.06 | Parkinsonism | Female |
| 01/01/2011 | 31/12/2011 | 634 | 2545789.566 | 24.9 | 23 | 26.92 | Parkinsonism | Female |
| 01/01/2012 | 31/12/2012 | 689 | 2529825.58 | 27.24 | 25.24 | 29.35 | Parkinsonism | Female |
| 01/01/2013 | 31/12/2013 | 681 | 2434753.098 | 27.97 | 25.91 | 30.15 | Parkinsonism | Female |
| 01/01/2014 | 31/12/2014 | 612 | 2266793.558 | 27 | 24.9 | 29.22 | Parkinsonism | Female |
| 01/01/2015 | 31/12/2015 | 545 | 2050972.082 | 26.57 | 24.39 | 28.9 | Parkinsonism | Female |
| 01/01/2016 | 31/12/2016 | 469 | 1779908.591 | 26.35 | 24.02 | 28.85 | Parkinsonism | Female |
| 01/01/2017 | 31/12/2017 | 423 | 1615385.64 | 26.19 | 23.75 | 28.8 | Parkinsonism | Female |
| 01/01/2018 | 31/12/2018 | 392 | 1534507.069 | 25.55 | 23.08 | 28.2 | Parkinsonism | Female |
| 01/01/2019 | 31/12/2019 | 395 | 1471420.085 | 26.84 | 24.26 | 29.63 | Parkinsonism | Female |
| 01/01/2020 | 31/12/2020 | 250 | 1372779.806 | 18.21 | 16.02 | 20.61 | Parkinsonism | Female |
| 01/01/2021 | 31/12/2021 | 273 | 1239716.843 | 22.02 | 19.49 | 24.79 | Parkinsonism | Female |
| 01/01/2007 | 31/12/2007 | 728 | 2541703.573 | 28.64 | 26.6 | 30.8 | ParkinsonsDisease | Female |
| 01/01/2008 | 31/12/2008 | 722 | 2586756.402 | 27.91 | 25.91 | 30.02 | ParkinsonsDisease | Female |
| 01/01/2009 | 31/12/2009 | 732 | 2601151.335 | 28.14 | 26.14 | 30.26 | ParkinsonsDisease | Female |
| 01/01/2010 | 31/12/2010 | 657 | 2583745.807 | 25.43 | 23.52 | 27.45 | ParkinsonsDisease | Female |
| 01/01/2011 | 31/12/2011 | 589 | 2546075.132 | 23.13 | 21.3 | 25.08 | ParkinsonsDisease | Female |
| 01/01/2012 | 31/12/2012 | 647 | 2530107.639 | 25.57 | 23.64 | 27.62 | ParkinsonsDisease | Female |
| 01/01/2013 | 31/12/2013 | 629 | 2435014.609 | 25.83 | 23.85 | 27.93 | ParkinsonsDisease | Female |
| 01/01/2014 | 31/12/2014 | 552 | 2267059.149 | 24.35 | 22.36 | 26.47 | ParkinsonsDisease | Female |
| 01/01/2015 | 31/12/2015 | 486 | 2051229.64 | 23.69 | 21.63 | 25.9 | ParkinsonsDisease | Female |
| 01/01/2016 | 31/12/2016 | 421 | 1780133.021 | 23.65 | 21.44 | 26.02 | ParkinsonsDisease | Female |
| 01/01/2017 | 31/12/2017 | 363 | 1615606.42 | 22.47 | 20.22 | 24.9 | ParkinsonsDisease | Female |
| 01/01/2018 | 31/12/2018 | 346 | 1534722.144 | 22.54 | 20.23 | 25.05 | ParkinsonsDisease | Female |
| 01/01/2019 | 31/12/2019 | 350 | 1471631.732 | 23.78 | 21.36 | 26.41 | ParkinsonsDisease | Female |
| 01/01/2020 | 31/12/2020 | 222 | 1372985.862 | 16.17 | 14.11 | 18.44 | ParkinsonsDisease | Female |
| 01/01/2021 | 31/12/2021 | 248 | 1239899.406 | 20 | 17.59 | 22.65 | ParkinsonsDisease | Female |
| 01/01/2007 | 31/12/2007 | 2 | 2545918.53 | 0.08 | 0.01 | 0.28 | VascularParkinsonism | Female |
| 01/01/2008 | 31/12/2008 | 1 | 2591082.921 | 0.04 | 0 | 0.22 | VascularParkinsonism | Female |
| 01/01/2009 | 31/12/2009 | 2 | 2605546.215 | 0.08 | 0.01 | 0.28 | VascularParkinsonism | Female |
| 01/01/2010 | 31/12/2010 | 30 | 2588105.413 | 1.16 | 0.78 | 1.65 | VascularParkinsonism | Female |
| 01/01/2011 | 31/12/2011 | 42 | 2550339.699 | 1.65 | 1.19 | 2.23 | VascularParkinsonism | Female |
| 01/01/2012 | 31/12/2012 | 40 | 2534326.119 | 1.58 | 1.13 | 2.15 | VascularParkinsonism | Female |
| 01/01/2013 | 31/12/2013 | 35 | 2439058.576 | 1.43 | 1 | 2 | VascularParkinsonism | Female |
| 01/01/2014 | 31/12/2014 | 41 | 2270837.539 | 1.81 | 1.3 | 2.45 | VascularParkinsonism | Female |
| 01/01/2015 | 31/12/2015 | 36 | 2054551.833 | 1.75 | 1.23 | 2.43 | VascularParkinsonism | Female |
| 01/01/2016 | 31/12/2016 | 41 | 1782917.717 | 2.3 | 1.65 | 3.12 | VascularParkinsonism | Female |
| 01/01/2017 | 31/12/2017 | 35 | 1618086.45 | 2.16 | 1.51 | 3.01 | VascularParkinsonism | Female |
| 01/01/2018 | 31/12/2018 | 20 | 1537075.535 | 1.3 | 0.79 | 2.01 | VascularParkinsonism | Female |
| 01/01/2019 | 31/12/2019 | 28 | 1473887.962 | 1.9 | 1.26 | 2.75 | VascularParkinsonism | Female |
| 01/01/2020 | 31/12/2020 | 17 | 1375008.03 | 1.24 | 0.72 | 1.98 | VascularParkinsonism | Female |
| 01/01/2021 | 31/12/2021 | 14 | 1241712.192 | 1.13 | 0.62 | 1.89 | VascularParkinsonism | Female |

Supplementary Table S4: Age-standardized prevalence of parkinsonism and subtypes and then stratified by sex

| prevalence_start_date | prevalence_end_date | n_events | person-years | standardised_prevalence | standardised_prevalence_lower | standardised_prevalence_upper | outcome_cohort_name | denominator_sex |
| --- | --- | --- | --- | --- | --- | --- | --- | --- |
| 01/01/2007 | 31/12/2007 | 140 | 5402479 | 2.59E-05 | 2.18E-05 | 3.06E-05 | DrugInducedParkinsonism | Both |
| 01/01/2008 | 31/12/2008 | 163 | 5463876 | 2.98E-05 | 2.54E-05 | 3.48E-05 | DrugInducedParkinsonism | Both |
| 01/01/2009 | 31/12/2009 | 160 | 5481298 | 2.92E-05 | 2.48E-05 | 3.41E-05 | DrugInducedParkinsonism | Both |
| 01/01/2010 | 31/12/2010 | 150 | 5488310 | 2.73E-05 | 2.31E-05 | 3.21E-05 | DrugInducedParkinsonism | Both |
| 01/01/2011 | 31/12/2011 | 120 | 5404852 | 2.22E-05 | 1.84E-05 | 2.65E-05 | DrugInducedParkinsonism | Both |
| 01/01/2012 | 31/12/2012 | 110 | 5315011 | 2.07E-05 | 1.70E-05 | 2.49E-05 | DrugInducedParkinsonism | Both |
| 01/01/2013 | 31/12/2013 | 111 | 5233690 | 2.12E-05 | 1.74E-05 | 2.55E-05 | DrugInducedParkinsonism | Both |
| 01/01/2014 | 31/12/2014 | 110 | 4957050 | 2.22E-05 | 1.82E-05 | 2.67E-05 | DrugInducedParkinsonism | Both |
| 01/01/2015 | 31/12/2015 | 108 | 4550988 | 2.37E-05 | 1.95E-05 | 2.87E-05 | DrugInducedParkinsonism | Both |
| 01/01/2016 | 31/12/2016 | 91 | 3895129 | 2.34E-05 | 1.88E-05 | 2.87E-05 | DrugInducedParkinsonism | Both |
| 01/01/2017 | 31/12/2017 | 76 | 3508465 | 2.17E-05 | 1.71E-05 | 2.71E-05 | DrugInducedParkinsonism | Both |
| 01/01/2018 | 31/12/2018 | 77 | 3271448 | 2.35E-05 | 1.86E-05 | 2.94E-05 | DrugInducedParkinsonism | Both |
| 01/01/2019 | 31/12/2019 | 91 | 3167317 | 2.87E-05 | 2.31E-05 | 3.53E-05 | DrugInducedParkinsonism | Both |
| 01/01/2020 | 31/12/2020 | 65 | 2985903 | 2.18E-05 | 1.68E-05 | 2.77E-05 | DrugInducedParkinsonism | Both |
| 01/01/2021 | 31/12/2021 | 56 | 2723881 | 2.06E-05 | 1.55E-05 | 2.67E-05 | DrugInducedParkinsonism | Both |
| 01/01/2007 | 31/12/2007 | 11934 | 5402479 | 0.002208986 | 0.002169529 | 0.00224898 | Parkinsonism | Both |
| 01/01/2008 | 31/12/2008 | 12390 | 5463876 | 0.002267621 | 0.002227866 | 0.002307907 | Parkinsonism | Both |
| 01/01/2009 | 31/12/2009 | 12753 | 5481298 | 0.002326639 | 0.002286431 | 0.002367376 | Parkinsonism | Both |
| 01/01/2010 | 31/12/2010 | 12850 | 5488310 | 0.00234134 | 0.002301031 | 0.002382178 | Parkinsonism | Both |
| 01/01/2011 | 31/12/2011 | 12761 | 5404852 | 0.002361027 | 0.002320238 | 0.002402353 | Parkinsonism | Both |
| 01/01/2012 | 31/12/2012 | 12818 | 5315011 | 0.00241166 | 0.002370089 | 0.002453778 | Parkinsonism | Both |
| 01/01/2013 | 31/12/2013 | 12687 | 5233690 | 0.002424102 | 0.002382102 | 0.002466657 | Parkinsonism | Both |
| 01/01/2014 | 31/12/2014 | 12203 | 4957050 | 0.002461746 | 0.00241826 | 0.002505818 | Parkinsonism | Both |
| 01/01/2015 | 31/12/2015 | 11237 | 4550988 | 0.002469134 | 0.00242369 | 0.002515217 | Parkinsonism | Both |
| 01/01/2016 | 31/12/2016 | 9672 | 3895129 | 0.002483101 | 0.002433859 | 0.00253309 | Parkinsonism | Both |
| 01/01/2017 | 31/12/2017 | 8518 | 3508465 | 0.002427842 | 0.002376554 | 0.002479958 | Parkinsonism | Both |
| 01/01/2018 | 31/12/2018 | 7945 | 3271448 | 0.002428588 | 0.002375477 | 0.002482588 | Parkinsonism | Both |
| 01/01/2019 | 31/12/2019 | 7843 | 3167317 | 0.002476228 | 0.002421726 | 0.002531648 | Parkinsonism | Both |
| 01/01/2020 | 31/12/2020 | 7110 | 2985903 | 0.002381189 | 0.002326158 | 0.002437193 | Parkinsonism | Both |
| 01/01/2021 | 31/12/2021 | 6423 | 2723881 | 0.002358033 | 0.002300714 | 0.002416418 | Parkinsonism | Both |
| 01/01/2007 | 31/12/2007 | 11432 | 5402479 | 0.002116066 | 0.002077452 | 0.002155217 | ParkinsonsDisease | Both |
| 01/01/2008 | 31/12/2008 | 11850 | 5463876 | 0.00216879 | 0.002129915 | 0.002208197 | ParkinsonsDisease | Both |
| 01/01/2009 | 31/12/2009 | 12166 | 5481298 | 0.002219547 | 0.00218028 | 0.002259344 | ParkinsonsDisease | Both |
| 01/01/2010 | 31/12/2010 | 12213 | 5488310 | 0.002225275 | 0.002185982 | 0.002265097 | ParkinsonsDisease | Both |
| 01/01/2011 | 31/12/2011 | 12087 | 5404852 | 0.002236324 | 0.002196632 | 0.002276554 | ParkinsonsDisease | Both |
| 01/01/2012 | 31/12/2012 | 12115 | 5315011 | 0.002279393 | 0.002238983 | 0.00232035 | ParkinsonsDisease | Both |
| 01/01/2013 | 31/12/2013 | 11961 | 5233690 | 0.002285386 | 0.00224461 | 0.002326716 | ParkinsonsDisease | Both |
| 01/01/2014 | 31/12/2014 | 11442 | 4957050 | 0.002308228 | 0.002266125 | 0.002350916 | ParkinsonsDisease | Both |
| 01/01/2015 | 31/12/2015 | 10503 | 4550988 | 0.002307851 | 0.002263922 | 0.002352417 | ParkinsonsDisease | Both |
| 01/01/2016 | 31/12/2016 | 9016 | 3895129 | 0.002314686 | 0.002267151 | 0.002362967 | ParkinsonsDisease | Both |
| 01/01/2017 | 31/12/2017 | 7905 | 3508465 | 0.002253122 | 0.002203724 | 0.002303348 | ParkinsonsDisease | Both |
| 01/01/2018 | 31/12/2018 | 7386 | 3271448 | 0.002257716 | 0.002206517 | 0.002309803 | ParkinsonsDisease | Both |
| 01/01/2019 | 31/12/2019 | 7263 | 3167317 | 0.002293108 | 0.002240671 | 0.002346463 | ParkinsonsDisease | Both |
| 01/01/2020 | 31/12/2020 | 6580 | 2985903 | 0.002203689 | 0.002150761 | 0.00225759 | ParkinsonsDisease | Both |
| 01/01/2021 | 31/12/2021 | 5916 | 2723881 | 0.002171901 | 0.002116905 | 0.002227964 | ParkinsonsDisease | Both |
| 01/01/2007 | 31/12/2007 | 6 | 5402479 | 1.11E-06 | 4.08E-07 | 2.42E-06 | VascularParkinsonism | Both |
| 01/01/2008 | 31/12/2008 | 9 | 5463876 | 1.65E-06 | 7.53E-07 | 3.13E-06 | VascularParkinsonism | Both |
| 01/01/2009 | 31/12/2009 | 18 | 5481298 | 3.28E-06 | 1.95E-06 | 5.19E-06 | VascularParkinsonism | Both |
| 01/01/2010 | 31/12/2010 | 96 | 5488310 | 1.75E-05 | 1.42E-05 | 2.14E-05 | VascularParkinsonism | Both |
| 01/01/2011 | 31/12/2011 | 199 | 5404852 | 3.68E-05 | 3.19E-05 | 4.23E-05 | VascularParkinsonism | Both |
| 01/01/2012 | 31/12/2012 | 290 | 5315011 | 5.46E-05 | 4.85E-05 | 6.12E-05 | VascularParkinsonism | Both |
| 01/01/2013 | 31/12/2013 | 352 | 5233690 | 6.73E-05 | 6.04E-05 | 7.47E-05 | VascularParkinsonism | Both |
| 01/01/2014 | 31/12/2014 | 402 | 4957050 | 8.11E-05 | 7.34E-05 | 8.94E-05 | VascularParkinsonism | Both |
| 01/01/2015 | 31/12/2015 | 427 | 4550988 | 9.38E-05 | 8.51E-05 | 0.000103162 | VascularParkinsonism | Both |
| 01/01/2016 | 31/12/2016 | 427 | 3895129 | 0.000109624 | 9.95E-05 | 0.000120532 | VascularParkinsonism | Both |
| 01/01/2017 | 31/12/2017 | 421 | 3508465 | 0.000119996 | 0.000108806 | 0.000132024 | VascularParkinsonism | Both |
| 01/01/2018 | 31/12/2018 | 369 | 3271448 | 0.000112794 | 0.000101578 | 0.000124911 | VascularParkinsonism | Both |
| 01/01/2019 | 31/12/2019 | 386 | 3167317 | 0.00012187 | 0.000110014 | 0.000134655 | VascularParkinsonism | Both |
| 01/01/2020 | 31/12/2020 | 348 | 2985903 | 0.000116548 | 0.000104623 | 0.000129459 | VascularParkinsonism | Both |
| 01/01/2021 | 31/12/2021 | 329 | 2723881 | 0.000120784 | 0.000108083 | 0.000134566 | VascularParkinsonism | Both |
| 01/01/2007 | 31/12/2007 | 50 | 2657280 | 1.88E-05 | 1.40E-05 | 2.48E-05 | DrugInducedParkinsonism | Male |
| 01/01/2008 | 31/12/2008 | 83 | 2689744 | 3.09E-05 | 2.46E-05 | 3.83E-05 | DrugInducedParkinsonism | Male |
| 01/01/2009 | 31/12/2009 | 71 | 2698962 | 2.63E-05 | 2.05E-05 | 3.32E-05 | DrugInducedParkinsonism | Male |
| 01/01/2010 | 31/12/2010 | 56 | 2703869 | 2.07E-05 | 1.56E-05 | 2.69E-05 | DrugInducedParkinsonism | Male |
| 01/01/2011 | 31/12/2011 | 58 | 2660965 | 2.18E-05 | 1.66E-05 | 2.82E-05 | DrugInducedParkinsonism | Male |
| 01/01/2012 | 31/12/2012 | 63 | 2614094 | 2.41E-05 | 1.85E-05 | 3.08E-05 | DrugInducedParkinsonism | Male |
| 01/01/2013 | 31/12/2013 | 63 | 2572677 | 2.45E-05 | 1.88E-05 | 3.13E-05 | DrugInducedParkinsonism | Male |
| 01/01/2014 | 31/12/2014 | 51 | 2434194 | 2.10E-05 | 1.56E-05 | 2.75E-05 | DrugInducedParkinsonism | Male |
| 01/01/2015 | 31/12/2015 | 44 | 2236771 | 1.97E-05 | 1.43E-05 | 2.64E-05 | DrugInducedParkinsonism | Male |
| 01/01/2016 | 31/12/2016 | 47 | 1914896 | 2.45E-05 | 1.80E-05 | 3.26E-05 | DrugInducedParkinsonism | Male |
| 01/01/2017 | 31/12/2017 | 37 | 1726463 | 2.14E-05 | 1.51E-05 | 2.95E-05 | DrugInducedParkinsonism | Male |
| 01/01/2018 | 31/12/2018 | 27 | 1611874 | 1.68E-05 | 1.10E-05 | 2.44E-05 | DrugInducedParkinsonism | Male |
| 01/01/2019 | 31/12/2019 | 34 | 1561971 | 2.18E-05 | 1.51E-05 | 3.04E-05 | DrugInducedParkinsonism | Male |
| 01/01/2020 | 31/12/2020 | 22 | 1473771 | 1.49E-05 | 9.36E-06 | 2.26E-05 | DrugInducedParkinsonism | Male |
| 01/01/2021 | 31/12/2021 | 21 | 1345357 | 1.56E-05 | 9.66E-06 | 2.39E-05 | DrugInducedParkinsonism | Male |
| 01/01/2007 | 31/12/2007 | 6647 | 2657280 | 0.00250143 | 0.002441653 | 0.002562301 | Parkinsonism | Male |
| 01/01/2008 | 31/12/2008 | 6969 | 2689744 | 0.002590953 | 0.002530475 | 0.002652511 | Parkinsonism | Male |
| 01/01/2009 | 31/12/2009 | 7229 | 2698962 | 0.002678437 | 0.002617045 | 0.002740906 | Parkinsonism | Male |
| 01/01/2010 | 31/12/2010 | 7340 | 2703869 | 0.002714629 | 0.002652877 | 0.002777455 | Parkinsonism | Male |
| 01/01/2011 | 31/12/2011 | 7390 | 2660965 | 0.002777188 | 0.002714226 | 0.002841242 | Parkinsonism | Male |
| 01/01/2012 | 31/12/2012 | 7470 | 2614094 | 0.002857587 | 0.002793148 | 0.002923137 | Parkinsonism | Male |
| 01/01/2013 | 31/12/2013 | 7409 | 2572677 | 0.00287988 | 0.002814673 | 0.002946216 | Parkinsonism | Male |
| 01/01/2014 | 31/12/2014 | 7186 | 2434194 | 0.002952107 | 0.002884241 | 0.003021166 | Parkinsonism | Male |
| 01/01/2015 | 31/12/2015 | 6657 | 2236771 | 0.002976165 | 0.002905096 | 0.003048534 | Parkinsonism | Male |
| 01/01/2016 | 31/12/2016 | 5818 | 1914896 | 0.003038285 | 0.00296071 | 0.003117379 | Parkinsonism | Male |
| 01/01/2017 | 31/12/2017 | 5108 | 1726463 | 0.00295865 | 0.002878064 | 0.003040921 | Parkinsonism | Male |
| 01/01/2018 | 31/12/2018 | 4782 | 1611874 | 0.002966733 | 0.002883237 | 0.003052034 | Parkinsonism | Male |
| 01/01/2019 | 31/12/2019 | 4764 | 1561971 | 0.003049993 | 0.002963992 | 0.003137855 | Parkinsonism | Male |
| 01/01/2020 | 31/12/2020 | 4326 | 1473771 | 0.002935327 | 0.002848501 | 0.003024127 | Parkinsonism | Male |
| 01/01/2021 | 31/12/2021 | 3938 | 1345357 | 0.002927104 | 0.002836389 | 0.003019983 | Parkinsonism | Male |
| 01/01/2007 | 31/12/2007 | 6451 | 2657280 | 0.00242767 | 0.002368786 | 0.002487648 | ParkinsonsDisease | Male |
| 01/01/2008 | 31/12/2008 | 6734 | 2689744 | 0.002503584 | 0.002444141 | 0.002564108 | ParkinsonsDisease | Male |
| 01/01/2009 | 31/12/2009 | 6963 | 2698962 | 0.002579881 | 0.002519636 | 0.002641203 | ParkinsonsDisease | Male |
| 01/01/2010 | 31/12/2010 | 7049 | 2703869 | 0.002607005 | 0.002546497 | 0.002668588 | ParkinsonsDisease | Male |
| 01/01/2011 | 31/12/2011 | 7054 | 2660965 | 0.002650918 | 0.002589412 | 0.002713516 | ParkinsonsDisease | Male |
| 01/01/2012 | 31/12/2012 | 7093 | 2614094 | 0.002713368 | 0.002650586 | 0.002777262 | ParkinsonsDisease | Male |
| 01/01/2013 | 31/12/2013 | 7001 | 2572677 | 0.00272129 | 0.002657914 | 0.002785795 | ParkinsonsDisease | Male |
| 01/01/2014 | 31/12/2014 | 6755 | 2434194 | 0.002775046 | 0.002709259 | 0.002842027 | ParkinsonsDisease | Male |
| 01/01/2015 | 31/12/2015 | 6249 | 2236771 | 0.002793759 | 0.002724916 | 0.002863902 | ParkinsonsDisease | Male |
| 01/01/2016 | 31/12/2016 | 5449 | 1914896 | 0.002845585 | 0.002770527 | 0.002922163 | ParkinsonsDisease | Male |
| 01/01/2017 | 31/12/2017 | 4774 | 1726463 | 0.002765191 | 0.002687302 | 0.002844765 | ParkinsonsDisease | Male |
| 01/01/2018 | 31/12/2018 | 4491 | 1611874 | 0.002786198 | 0.0027053 | 0.0028689 | ParkinsonsDisease | Male |
| 01/01/2019 | 31/12/2019 | 4442 | 1561971 | 0.002843843 | 0.00276082 | 0.002928728 | ParkinsonsDisease | Male |
| 01/01/2020 | 31/12/2020 | 4038 | 1473771 | 0.00273991 | 0.002656046 | 0.002825749 | ParkinsonsDisease | Male |
| 01/01/2021 | 31/12/2021 | 3652 | 1345357 | 0.002714521 | 0.002627188 | 0.002804018 | ParkinsonsDisease | Male |
| 01/01/2007 | 31/12/2007 | 2 | 2657280 | 7.53E-07 | 9.11E-08 | 2.72E-06 | VascularParkinsonism | Male |
| 01/01/2008 | 31/12/2008 | 4 | 2689744 | 1.49E-06 | 4.05E-07 | 3.81E-06 | VascularParkinsonism | Male |
| 01/01/2009 | 31/12/2009 | 11 | 2698962 | 4.08E-06 | 2.03E-06 | 7.29E-06 | VascularParkinsonism | Male |
| 01/01/2010 | 31/12/2010 | 58 | 2703869 | 2.15E-05 | 1.63E-05 | 2.77E-05 | VascularParkinsonism | Male |
| 01/01/2011 | 31/12/2011 | 117 | 2660965 | 4.40E-05 | 3.64E-05 | 5.27E-05 | VascularParkinsonism | Male |
| 01/01/2012 | 31/12/2012 | 179 | 2614094 | 6.85E-05 | 5.88E-05 | 7.93E-05 | VascularParkinsonism | Male |
| 01/01/2013 | 31/12/2013 | 231 | 2572677 | 8.98E-05 | 7.86E-05 | 0.000102146 | VascularParkinsonism | Male |
| 01/01/2014 | 31/12/2014 | 262 | 2434194 | 0.000107633 | 9.50E-05 | 0.000121486 | VascularParkinsonism | Male |
| 01/01/2015 | 31/12/2015 | 275 | 2236771 | 0.000122945 | 0.000108842 | 0.000138368 | VascularParkinsonism | Male |
| 01/01/2016 | 31/12/2016 | 266 | 1914896 | 0.000138911 | 0.000122718 | 0.000156647 | VascularParkinsonism | Male |
| 01/01/2017 | 31/12/2017 | 251 | 1726463 | 0.000145384 | 0.000127953 | 0.000164527 | VascularParkinsonism | Male |
| 01/01/2018 | 31/12/2018 | 221 | 1611874 | 0.000137108 | 0.000119626 | 0.000156426 | VascularParkinsonism | Male |
| 01/01/2019 | 31/12/2019 | 247 | 1561971 | 0.000158134 | 0.000139026 | 0.000179134 | VascularParkinsonism | Male |
| 01/01/2020 | 31/12/2020 | 218 | 1473771 | 0.00014792 | 0.000128935 | 0.000168914 | VascularParkinsonism | Male |
| 01/01/2021 | 31/12/2021 | 214 | 1345357 | 0.000159066 | 0.000138466 | 0.000181865 | VascularParkinsonism | Male |
| 01/01/2007 | 31/12/2007 | 90 | 2745199 | 3.28E-05 | 2.64E-05 | 4.03E-05 | DrugInducedParkinsonism | Female |
| 01/01/2008 | 31/12/2008 | 80 | 2774132 | 2.88E-05 | 2.29E-05 | 3.59E-05 | DrugInducedParkinsonism | Female |
| 01/01/2009 | 31/12/2009 | 89 | 2782336 | 3.20E-05 | 2.57E-05 | 3.94E-05 | DrugInducedParkinsonism | Female |
| 01/01/2010 | 31/12/2010 | 94 | 2784441 | 3.38E-05 | 2.73E-05 | 4.13E-05 | DrugInducedParkinsonism | Female |
| 01/01/2011 | 31/12/2011 | 62 | 2743887 | 2.26E-05 | 1.73E-05 | 2.90E-05 | DrugInducedParkinsonism | Female |
| 01/01/2012 | 31/12/2012 | 47 | 2700917 | 1.74E-05 | 1.28E-05 | 2.31E-05 | DrugInducedParkinsonism | Female |
| 01/01/2013 | 31/12/2013 | 48 | 2661013 | 1.80E-05 | 1.33E-05 | 2.39E-05 | DrugInducedParkinsonism | Female |
| 01/01/2014 | 31/12/2014 | 59 | 2522856 | 2.34E-05 | 1.78E-05 | 3.02E-05 | DrugInducedParkinsonism | Female |
| 01/01/2015 | 31/12/2015 | 64 | 2314217 | 2.77E-05 | 2.13E-05 | 3.53E-05 | DrugInducedParkinsonism | Female |
| 01/01/2016 | 31/12/2016 | 44 | 1980233 | 2.22E-05 | 1.61E-05 | 2.98E-05 | DrugInducedParkinsonism | Female |
| 01/01/2017 | 31/12/2017 | 39 | 1782002 | 2.19E-05 | 1.56E-05 | 2.99E-05 | DrugInducedParkinsonism | Female |
| 01/01/2018 | 31/12/2018 | 50 | 1659574 | 3.01E-05 | 2.24E-05 | 3.97E-05 | DrugInducedParkinsonism | Female |
| 01/01/2019 | 31/12/2019 | 57 | 1605346 | 3.55E-05 | 2.69E-05 | 4.60E-05 | DrugInducedParkinsonism | Female |
| 01/01/2020 | 31/12/2020 | 43 | 1512132 | 2.84E-05 | 2.06E-05 | 3.83E-05 | DrugInducedParkinsonism | Female |
| 01/01/2021 | 31/12/2021 | 35 | 1378524 | 2.54E-05 | 1.77E-05 | 3.53E-05 | DrugInducedParkinsonism | Female |
| 01/01/2007 | 31/12/2007 | 5287 | 2745199 | 0.001925908 | 0.00187434 | 0.001978534 | Parkinsonism | Female |
| 01/01/2008 | 31/12/2008 | 5421 | 2774132 | 0.001954125 | 0.001902448 | 0.002006849 | Parkinsonism | Female |
| 01/01/2009 | 31/12/2009 | 5524 | 2782336 | 0.001985382 | 0.001933367 | 0.002038442 | Parkinsonism | Female |
| 01/01/2010 | 31/12/2010 | 5510 | 2784441 | 0.001978853 | 0.001926944 | 0.002031806 | Parkinsonism | Female |
| 01/01/2011 | 31/12/2011 | 5371 | 2743887 | 0.001957442 | 0.001905439 | 0.002010505 | Parkinsonism | Female |
| 01/01/2012 | 31/12/2012 | 5348 | 2700917 | 0.001980068 | 0.001927352 | 0.002033861 | Parkinsonism | Female |
| 01/01/2013 | 31/12/2013 | 5278 | 2661013 | 0.001983455 | 0.001930302 | 0.002037701 | Parkinsonism | Female |
| 01/01/2014 | 31/12/2014 | 5017 | 2522856 | 0.001988619 | 0.001933968 | 0.002044423 | Parkinsonism | Female |
| 01/01/2015 | 31/12/2015 | 4580 | 2314217 | 0.001979071 | 0.001922165 | 0.002037234 | Parkinsonism | Female |
| 01/01/2016 | 31/12/2016 | 3854 | 1980233 | 0.001946236 | 0.00188527 | 0.002008671 | Parkinsonism | Female |
| 01/01/2017 | 31/12/2017 | 3410 | 1782002 | 0.001913578 | 0.001849884 | 0.001978905 | Parkinsonism | Female |
| 01/01/2018 | 31/12/2018 | 3163 | 1659574 | 0.001905911 | 0.001840063 | 0.001973513 | Parkinsonism | Female |
| 01/01/2019 | 31/12/2019 | 3079 | 1605346 | 0.001917967 | 0.001850812 | 0.001986935 | Parkinsonism | Female |
| 01/01/2020 | 31/12/2020 | 2784 | 1512132 | 0.001841109 | 0.001773348 | 0.001910797 | Parkinsonism | Female |
| 01/01/2021 | 31/12/2021 | 2485 | 1378524 | 0.001802653 | 0.001732467 | 0.001874953 | Parkinsonism | Female |
| 01/01/2007 | 31/12/2007 | 4981 | 2745199 | 0.00181444 | 0.001764398 | 0.001865543 | ParkinsonsDisease | Female |
| 01/01/2008 | 31/12/2008 | 5116 | 2774132 | 0.00184418 | 0.001793988 | 0.001895421 | ParkinsonsDisease | Female |
| 01/01/2009 | 31/12/2009 | 5203 | 2782336 | 0.001870011 | 0.001819541 | 0.001921527 | ParkinsonsDisease | Female |
| 01/01/2010 | 31/12/2010 | 5164 | 2784441 | 0.001854591 | 0.00180435 | 0.001905877 | ParkinsonsDisease | Female |
| 01/01/2011 | 31/12/2011 | 5033 | 2743887 | 0.001834259 | 0.00178393 | 0.001885648 | ParkinsonsDisease | Female |
| 01/01/2012 | 31/12/2012 | 5022 | 2700917 | 0.001859369 | 0.001808295 | 0.001911519 | ParkinsonsDisease | Female |
| 01/01/2013 | 31/12/2013 | 4960 | 2661013 | 0.001863952 | 0.001812436 | 0.001916561 | ParkinsonsDisease | Female |
| 01/01/2014 | 31/12/2014 | 4687 | 2522856 | 0.001857815 | 0.001805005 | 0.001911778 | ParkinsonsDisease | Female |
| 01/01/2015 | 31/12/2015 | 4254 | 2314217 | 0.001838203 | 0.001783375 | 0.001894288 | ParkinsonsDisease | Female |
| 01/01/2016 | 31/12/2016 | 3567 | 1980233 | 0.001801303 | 0.00174267 | 0.001861406 | ParkinsonsDisease | Female |
| 01/01/2017 | 31/12/2017 | 3131 | 1782002 | 0.001757013 | 0.001696002 | 0.001819657 | ParkinsonsDisease | Female |
| 01/01/2018 | 31/12/2018 | 2895 | 1659574 | 0.001744424 | 0.001681452 | 0.00180915 | ParkinsonsDisease | Female |
| 01/01/2019 | 31/12/2019 | 2821 | 1605346 | 0.001757254 | 0.001693 | 0.001823322 | ParkinsonsDisease | Female |
| 01/01/2020 | 31/12/2020 | 2542 | 1512132 | 0.00168107 | 0.001616349 | 0.001747719 | ParkinsonsDisease | Female |
| 01/01/2021 | 31/12/2021 | 2264 | 1378524 | 0.001642336 | 0.001575375 | 0.001711412 | ParkinsonsDisease | Female |
| 01/01/2007 | 31/12/2007 | 4 | 2745199 | 1.46E-06 | 3.97E-07 | 3.73E-06 | VascularParkinsonism | Female |
| 01/01/2008 | 31/12/2008 | 5 | 2774132 | 1.80E-06 | 5.85E-07 | 4.21E-06 | VascularParkinsonism | Female |
| 01/01/2009 | 31/12/2009 | 7 | 2782336 | 2.52E-06 | 1.01E-06 | 5.18E-06 | VascularParkinsonism | Female |
| 01/01/2010 | 31/12/2010 | 38 | 2784441 | 1.36E-05 | 9.66E-06 | 1.87E-05 | VascularParkinsonism | Female |
| 01/01/2011 | 31/12/2011 | 82 | 2743887 | 2.99E-05 | 2.38E-05 | 3.71E-05 | VascularParkinsonism | Female |
| 01/01/2012 | 31/12/2012 | 111 | 2700917 | 4.11E-05 | 3.38E-05 | 4.95E-05 | VascularParkinsonism | Female |
| 01/01/2013 | 31/12/2013 | 121 | 2661013 | 4.55E-05 | 3.77E-05 | 5.43E-05 | VascularParkinsonism | Female |
| 01/01/2014 | 31/12/2014 | 140 | 2522856 | 5.55E-05 | 4.67E-05 | 6.55E-05 | VascularParkinsonism | Female |
| 01/01/2015 | 31/12/2015 | 152 | 2314217 | 6.57E-05 | 5.57E-05 | 7.70E-05 | VascularParkinsonism | Female |
| 01/01/2016 | 31/12/2016 | 161 | 1980233 | 8.13E-05 | 6.92E-05 | 9.49E-05 | VascularParkinsonism | Female |
| 01/01/2017 | 31/12/2017 | 170 | 1782002 | 9.54E-05 | 8.16E-05 | 0.000110866 | VascularParkinsonism | Female |
| 01/01/2018 | 31/12/2018 | 148 | 1659574 | 8.92E-05 | 7.54E-05 | 0.00010476 | VascularParkinsonism | Female |
| 01/01/2019 | 31/12/2019 | 139 | 1605346 | 8.66E-05 | 7.28E-05 | 0.000102235 | VascularParkinsonism | Female |
| 01/01/2020 | 31/12/2020 | 130 | 1512132 | 8.60E-05 | 7.18E-05 | 0.000102084 | VascularParkinsonism | Female |
| 01/01/2021 | 31/12/2021 | 115 | 1378524 | 8.34E-05 | 6.89E-05 | 0.000100136 | VascularParkinsonism | Female |
